# Evaluation of individual and ensemble probabilistic forecasts of COVID-19 mortality in the US

**DOI:** 10.1101/2021.02.03.21250974

**Authors:** Estee Y Cramer, Evan L Ray, Velma K Lopez, Johannes Bracher, Andrea Brennen, Alvaro J Castro Rivadeneira, Aaron Gerding, Tilmann Gneiting, Katie H House, Yuxin Huang, Dasuni Jayawardena, Abdul H Kanji, Ayush Khandelwal, Khoa Le, Anja Mühlemann, Jarad Niemi, Apurv Shah, Ariane Stark, Yijin Wang, Nutcha Wattanachit, Martha W Zorn, Youyang Gu, Sansiddh Jain, Nayana Bannur, Ayush Deva, Mihir Kulkarni, Srujana Merugu, Alpan Raval, Siddhant Shingi, Avtansh Tiwari, Jerome White, Neil F Abernethy, Spencer Woody, Maytal Dahan, Spencer Fox, Kelly Gaither, Michael Lachmann, Lauren Ancel Meyers, James G Scott, Mauricio Tec, Ajitesh Srivastava, Glover E George, Jeffrey C Cegan, Ian D Dettwiller, William P England, Matthew W Farthing, Robert H Hunter, Brandon Lafferty, Igor Linkov, Michael L Mayo, Matthew D Parno, Michael A Rowland, Benjamin D Trump, Yanli Zhang-James, Samuel Chen, Stephen V Faraone, Jonathan Hess, Christopher P Morley, Asif Salekin, Dongliang Wang, Sabrina M Corsetti, Thomas M Baer, Marisa C Eisenberg, Karl Falb, Yitao Huang, Emily T Martin, Ella McCauley, Robert L Myers, Tom Schwarz, Daniel Sheldon, Graham Casey Gibson, Rose Yu, Liyao Gao, Yian Ma, Dongxia Wu, Xifeng Yan, Xiaoyong Jin, Yu-Xiang Wang, YangQuan Chen, Lihong Guo, Yanting Zhao, Quanquan Gu, Jinghui Chen, Lingxiao Wang, Pan Xu, Weitong Zhang, Difan Zou, Hannah Biegel, Joceline Lega, Steve McConnell, VP Nagraj, Stephanie L Guertin, Christopher Hulme-Lowe, Stephen D Turner, Yunfeng Shi, Xuegang Ban, Robert Walraven, Qi-Jun Hong, Stanley Kong, Axel van de Walle, James A Turtle, Michal Ben-Nun, Steven Riley, Pete Riley, Ugur Koyluoglu, David DesRoches, Pedro Forli, Bruce Hamory, Christina Kyriakides, Helen Leis, John Milliken, Michael Moloney, James Morgan, Ninad Nirgudkar, Gokce Ozcan, Noah Piwonka, Matt Ravi, Chris Schrader, Elizabeth Shakhnovich, Daniel Siegel, Ryan Spatz, Chris Stiefeling, Barrie Wilkinson, Alexander Wong, Sean Cavany, Guido España, Sean Moore, Rachel Oidtman, Alex Perkins, David Kraus, Andrea Kraus, Zhifeng Gao, Jiang Bian, Wei Cao, Juan Lavista Ferres, Chaozhuo Li, Tie-Yan Liu, Xing Xie, Shun Zhang, Shun Zheng, Alessandro Vespignani, Matteo Chinazzi, Jessica T Davis, Kunpeng Mu, Ana Pastore y Piontti, Xinyue Xiong, Andrew Zheng, Jackie Baek, Vivek Farias, Andreea Georgescu, Retsef Levi, Deeksha Sinha, Joshua Wilde, Georgia Perakis, Mohammed Amine Bennouna, David Nze-Ndong, Divya Singhvi, Ioannis Spantidakis, Leann Thayaparan, Asterios Tsiourvas, Arnab Sarker, Ali Jadbabaie, Devavrat Shah, Nicolas Della Penna, Leo A Celi, Saketh Sundar, Russ Wolfinger, Dave Osthus, Lauren Castro, Geoffrey Fairchild, Isaac Michaud, Dean Karlen, Matt Kinsey, Luke C. Mullany, Kaitlin Rainwater-Lovett, Lauren Shin, Katharine Tallaksen, Shelby Wilson, Elizabeth C Lee, Juan Dent, Kyra H Grantz, Alison L Hill, Joshua Kaminsky, Kathryn Kaminsky, Lindsay T Keegan, Stephen A Lauer, Joseph C Lemaitre, Justin Lessler, Hannah R Meredith, Javier Perez-Saez, Sam Shah, Claire P Smith, Shaun A Truelove, Josh Wills, Maximilian Marshall, Lauren Gardner, Kristen Nixon, John C. Burant, Lily Wang, Lei Gao, Zhiling Gu, Myungjin Kim, Xinyi Li, Guannan Wang, Yueying Wang, Shan Yu, Robert C Reiner, Ryan Barber, Emmanuela Gakidou, Simon I. Hay, Steve Lim, Chris J.L. Murray, David Pigott, Heidi L Gurung, Prasith Baccam, Steven A Stage, Bradley T Suchoski, B. Aditya Prakash, Bijaya Adhikari, Jiaming Cui, Alexander Rodríguez, Anika Tabassum, Jiajia Xie, Pinar Keskinocak, John Asplund, Arden Baxter, Buse Eylul Oruc, Nicoleta Serban, Sercan O Arik, Mike Dusenberry, Arkady Epshteyn, Elli Kanal, Long T Le, Chun-Liang Li, Tomas Pfister, Dario Sava, Rajarishi Sinha, Thomas Tsai, Nate Yoder, Jinsung Yoon, Leyou Zhang, Sam Abbott, Nikos I Bosse, Sebastian Funk, Joel Hellewell, Sophie R Meakin, Katharine Sherratt, Mingyuan Zhou, Rahi Kalantari, Teresa K Yamana, Sen Pei, Jeffrey Shaman, Michael L Li, Dimitris Bertsimas, Omar Skali Lami, Saksham Soni, Hamza Tazi Bouardi, Turgay Ayer, Madeline Adee, Jagpreet Chhatwal, Ozden O Dalgic, Mary A Ladd, Benjamin P Linas, Peter Mueller, Jade Xiao, Yuanjia Wang, Qinxia Wang, Shanghong Xie, Donglin Zeng, Alden Green, Jacob Bien, Logan Brooks, Addison J Hu, Maria Jahja, Daniel McDonald, Balasubramanian Narasimhan, Collin Politsch, Samyak Rajanala, Aaron Rumack, Noah Simon, Ryan J Tibshirani, Rob Tibshirani, Valerie Ventura, Larry Wasserman, Eamon B O’Dea, John M Drake, Robert Pagano, Quoc T Tran, Lam Si Tung Ho, Huong Huynh, Jo W Walker, Rachel B Slayton, Michael A Johansson, Matthew Biggerstaff, Nicholas G Reich

## Abstract

Short-term probabilistic forecasts of the trajectory of the COVID-19 pandemic in the United States have served as a visible and important communication channel between the scientific modeling community and both the general public and decision-makers. Forecasting models provide specific, quantitative, and evaluable predictions that inform short-term decisions such as healthcare staffing needs, school closures, and allocation of medical supplies. Starting in April 2020, the US COVID-19 Forecast Hub (https://covid19forecasthub.org/) collected, disseminated, and synthesized tens of millions of specific predictions from more than 90 different academic, industry, and independent research groups. A multi-model ensemble forecast that combined predictions from dozens of different research groups every week provided the most consistently accurate probabilistic forecasts of incident deaths due to COVID-19 at the state and national level from April 2020 through October 2021. The performance of 27 individual models that submitted complete forecasts of COVID-19 deaths consistently throughout this year showed high variability in forecast skill across time, geospatial units, and forecast horizons. Two-thirds of the models evaluated showed better accuracy than a naïve baseline model. Forecast accuracy degraded as models made predictions further into the future, with probabilistic error at a 20-week horizon 3-5 times larger than when predicting at a 1-week horizon. This project underscores the role that collaboration and active coordination between governmental public health agencies, academic modeling teams, and industry partners can play in developing modern modeling capabilities to support local, state, and federal response to outbreaks.

**Significance Statement:** This paper compares the probabilistic accuracy of short-term forecasts of reported deaths due to COVID-19 during the first year and a half of the pandemic in the US. Results show high variation in accuracy between and within stand-alone models, and more consistent accuracy from an ensemble model that combined forecasts from all eligible models. This demonstrates that an ensemble model provided a reliable and comparatively accurate means of forecasting deaths during the COVID-19 pandemic that exceeded the performance of all of the models that contributed to it. This work strengthens the evidence base for synthesizing multiple models to support public health action.

## Introduction

Effective pandemic response requires federal, state, and local leaders to make timely decisions in order to reduce disease transmission. During the COVID-19 pandemic, surveillance data on the number of cases, hospitalizations, and disease-associated deaths were used to inform response policies (1, 2). While these data provide insight into recent trends in the outbreak, they only present a partial, time-lagged picture of transmission and do not show if and when changes may occur in the future.

Anticipating outbreak change is critical for optimal resource allocation and response. Forecasting models provide quantitative, evaluable, and probabilistic predictions about the epidemic trajectory for the near-term future. Forecasts can inform operational decisions about allocation of healthcare supplies (e.g., personal protective equipment, therapeutics, and vaccines), staffing needs, and school closures (3). Providing prediction uncertainty is critical for such decisions, as it allows stakeholders to assess the most likely outcomes and plausible worst-case scenarios (3).

Academic research groups, government agencies, industry teams, and individuals produced COVID-19 forecasts at an unprecedented scale starting in March 2020. Publicly available forecasts reflect varied approaches, data sources, and assumptions. Some models had mechanisms that allowed them to incorporate an estimated impact of current or potential future policies on human behavior and COVID-19 transmission. Other models assumed that currently observed trends would continue into the future without considering external data on policies in different jurisdictions.

To leverage these forecasts for the COVID-19 response, the United States Centers for Disease Control and Prevention (CDC) partnered with the Reich Lab at the University of Massachusetts Amherst to create the COVID-19 Forecast Hub (https://covid19forecasthub.org/) (4). Launched in early April 2020, the Forecast Hub facilitated the collection, archiving, evaluation, and synthesis of forecasts. Teams were explicitly asked to submit “unconditional” forecasts of the future, in other words, predictions that integrate across all possible changes in future dynamics. In practice, most individual models make predictions that are conditional on explicit or implicit assumptions of how policies, behaviors, and pathogens will evolve in the coming weeks. From these forecasts, a multi-model ensemble was developed, published weekly in real-time, and used by CDC in official public communications about the pandemic (https://www.cdc.gov/coronavirus/2019-ncov/covid-data/mathematical-modeling.html). Forecasts were generated for the outcomes of reported cases, hospitalizations and deaths due to COVID-19. This paper focuses on evaluating forecasts of reported deaths.

Ensemble models incorporate the information and uncertainties from multiple forecasts, each with their own perspectives, strengths and limitations, to create accurate predictions with well-calibrated uncertainty (5–10). Synthesizing multiple models removes the risk of over-reliance on any single approach for accuracy or stability. It is challenging for individual models to make calibrated predictions of the future when the behavior of the system being studied is non-stationary due to continually changing policies and behaviors. Ensemble approaches have previously demonstrated superior performance compared with single models in forecasting influenza (11–13), Ebola (14), and dengue fever outbreaks (15). Preliminary research suggested that COVID-19 ensemble forecasts were also more accurate and precise than individual models in the early phases of the pandemic (16, 17).

Predicting the trajectory of a novel pathogen outbreak such as COVID-19 is subject to many challenges. These include the role of human behavior and decision-making in outbreak trajectories; the fact that epidemic forecasts may play a role in a “feedback loop” when and if the forecasts themselves have the ability to impact future societal or individual decision-making (18). There are also a host of data irregularities, especially in the early stages of the pandemic.

It is important to systematically and rigorously evaluate forecasts designed to predict real-time changes to the outbreak in order to identify strengths and weaknesses of different approaches and to understand the extent to which the forecasts are a reliable input to public health decisions. Knowledge of what leads to more or less accurate and well-calibrated forecasts can inform their development and their use within outbreak science and public policy. In this analysis, we sought to evaluate the accuracy of individual and ensemble probabilistic forecasts submitted to the Forecast Hub, focusing on forecasts of reported weekly incident deaths.

## Results

### Summary of models

Forecasts evaluated in this analysis are based on submissions in a continuous 79-week period starting late April 2020 and ending in late October 2021 (Figure 1, Methods). Forecasts were evaluated at 55 locations including all 50 states, 4 jurisdictions and territories (Guam, US Virgin Islands, Puerto Rico, and the District of Columbia), and the US national level. The evaluation period captured the decline of the spring 2020 wave, a late summer 2020 increase in several locations, a large late-fall/early-winter surge in 2020/2021, and the rise and fall of the Delta variant in the summer and fall of 2021 (Figure 1B).

**Figure 1:**
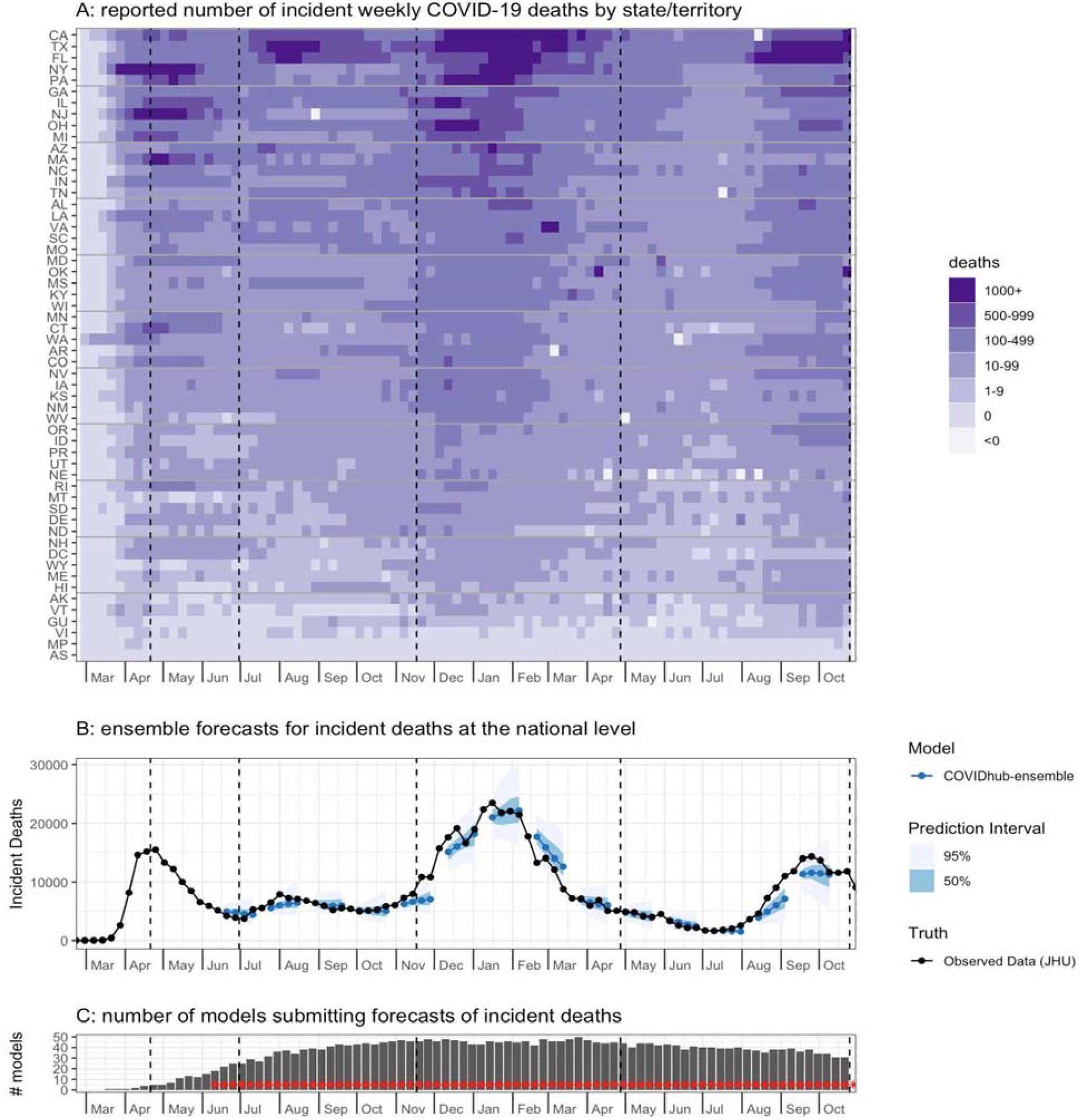
Overview of the evaluation period included in the paper. Vertical dashed lines indicate “phases’ ‘ of the pandemic analyzed separately in the supplement. (A) The reported number of incident weekly COVID-19 deaths by state or territory, per JHU CSSE reports. Locations are sorted by the cumulative number of deaths as of October 30th, 2021. (B) The time-series of weekly incident deaths at the national level overlaid with example forecasts from the COVID-19 Forecast Hub ensemble model. (C) The number of models submitting forecasts for incident deaths each week. Weeks in which the ensemble was submitted are shown with a red asterisk.

The number of models that submitted forecasts of incident deaths to the Forecast Hub and were screened for inclusion in this analysis increased from 4 models at the beginning of the evaluation period to an average of 41.2 models per week during the first ten months of 2021 (Figure 1C, Figure S1). Twenty-eight models met inclusion criteria, yielding 1,791 submission files with 556,050 specific predictions for unique combinations of forecast dates, targets (horizons forecasted), and locations.

The evaluated forecasts used different data sources and made varying assumptions about future transmission patterns (Table S1). All evaluated models other than CEID-Walk, the COVIDhub-baseline, the COVIDhub-ensemble, and PSI-Draft used case data as inputs to their forecast models. Ten models included data on COVID-19 hospitalizations, ten models incorporated demographic data, and nine models used mobility data. Of the 28 evaluated models, seven made explicit assumptions that social distancing and other behavioral patterns would change over the prediction period. Two naive models were included. The COVIDhub-baseline is a neutral model built with median predicted incidence equal to the number of reported deaths in the most recent week with uncertainty around the median based on weekly differences in previous observations (see Methods). CEID-Walk is a random walk model with simple outlier handling (Table S1).

### Overall model accuracy

To evaluate probabilistic accuracy, the primary metric used was the weighted interval score (WIS), a non-negative metric which measures how consistent a collection of prediction intervals is with an observed value (19). For WIS, a lower value represents smaller error (see Methods, SI Text).

Led by the ensemble model, a majority of the evaluated models achieved better accuracy than the baseline model in forecasting incident deaths (Table 1). The COVIDhub-ensemble achieved a relative WIS of 0.61, which can be interpreted as achieving, on average, 39% less probabilistic error than the baseline forecast in the evaluation period, adjusting for the ease or difficulty of the specific predictions made. An additional seven models achieved a relative WIS of less than or equal to 0.75. In total, 18 models had a relative WIS of less than 1, indicating lower probabilistic forecast error than the baseline model, and 10 models (including the baseline) had a relative WIS of 1 or greater (Table 1). Patterns in relative point forecast error were similar, with 18 models having equal or lower mean absolute error (MAE) than the baseline (Table 1). Values of relative WIS and rankings of models were robust to changing thresholds for submission inclusion criteria and to the inclusion or exclusion of individual outlying or revised observations (Tables S3 and S4). When stratified by phase of the pandemic, different models showed the highest accuracy overall (Figure S5).

**Table 1:**
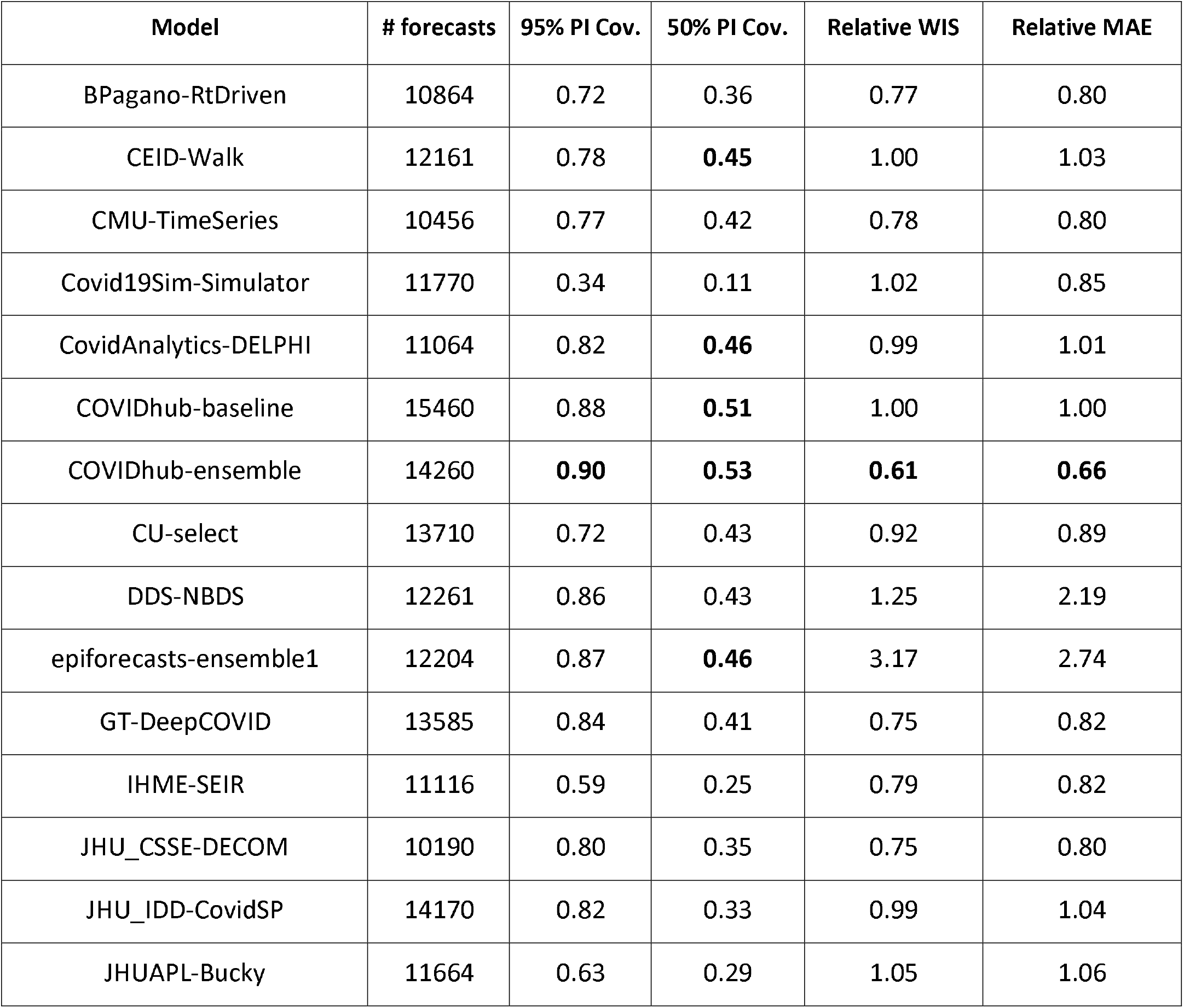
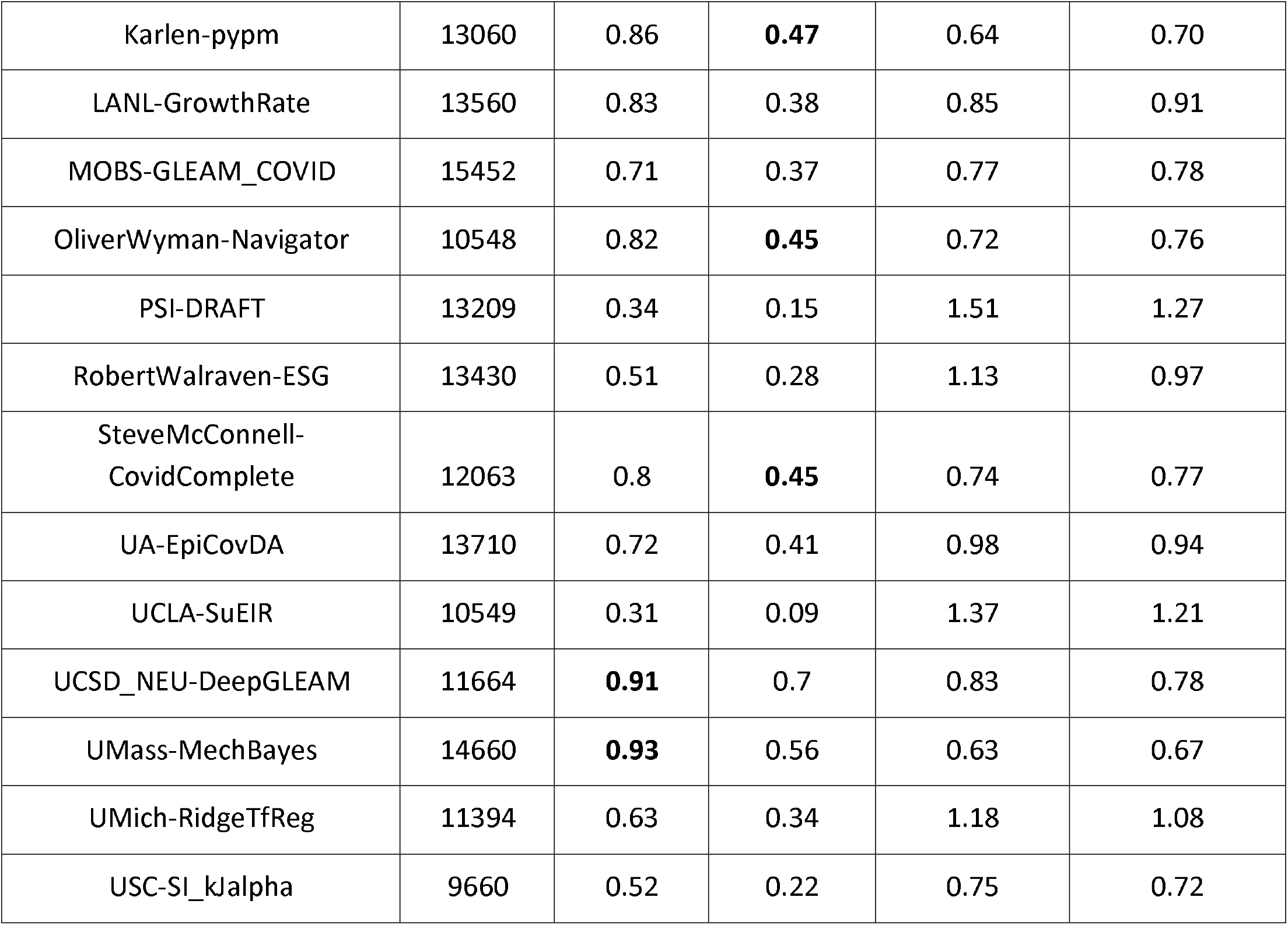
Summary accuracy metrics for all submitted forecasts from 28 models meeting inclusion criteria, aggregated across locations (50 states only), submission week, and 1- through 4-week forecast horizons. The ‘# forecasts’ column refers to the number of individual location-target-week combinations. Empirical prediction interval (PI) coverage rates calculate the fraction of times the 50% or 95% PIs covered the eventually observed value. Values within 5% coverage of the nominal rates are highlighted in boldface text. The “relative WIS” and “relative MAE” columns show the relative mean weighted interval score (WIS) and relative mean absolute error (MAE), which compare each model to the baseline model while adjusting for the difficulty of the forecasts the given model made for state-level forecasts (see Methods). The baseline model is defined to have a relative score of 1. Models with relative WIS or MAE values lower than 1 had “better” accuracy relative to the baseline model (best score in bold).

The degree to which individual models provided calibrated predictions varied widely (Table 1). We measured the probabilistic calibration of model forecasts using the empirical coverage rates of prediction intervals (PIs). Across 1 through 4 week-ahead horizons, 79 weeks, and 50 states, only the ensemble model achieved near nominal coverage rates for both the 50% and 95% PIs. Eight models achieved coverage rates within 5% of the desired coverage level for the 50% PI, and only the COVIDhub-ensemble and UMass-MechBayes achieved coverage rates within 5% for the 95% PI. Typically, observed coverage rates were lower than the nominal rate (Table 1, Figure S2). Three models had very low coverage rates (less than 50% for the 95% PI or less than 15% for the 50% PI). In general, models were penalized more for underpredicting the eventually observed values than overpredicting (Figure S7).

Among the top performing models, there was variation in data sources used, indicating that the inclusion of additional data sources was not a sufficient condition for high accuracy. Of the top individual models with a relative WIS less than or equal to 0.75, (UMass-MechBayes, Karlen-pypm, OliverWyman-Navigator, SteveMcConnell-CovidComplete, GT-DeepCovid, JHU_CSSE-DECOM, and USC-SI_kJalpha) four used data beyond the epidemiological hospitalization, case, and death surveillance data from CSSE (Table S1). Ten of the 18 individual models that performed better than the baseline used data other than epidemiological surveillance data (e.g., demographics or mobility). The top performers consisted of both models with mechanistic components and mostly phenomenological ones.

### Model accuracy rankings are highly variable

The COVIDhub-ensemble was the only model that ranked in the top half of all models (standardized rank > 0.5) for more than 85% of the observations it forecasted, although it made the single best forecast less frequently than any other model (Figure 2). We ranked models based on relative WIS for each combination of 1 through 4 week-ahead horizons, 79 weeks, and 55 locations, contributing to 17,006 possible predicted observations for each model (Figure 2). All models showed large variability in relative skill, with each model having observations for which it had the best (lowest) WIS and thereby a standardized rank of 1. Some models such as JHUAPL-BUCKY and PSI-DRAFT show a bimodal distribution of standardized rank, with one mode in the top quartile of models and another in the bottom quartile. In these cases, the models frequently made overconfident predictions (Figure S6) resulting in either lower scores (indicating better performance) in instances when their predictions were very close to the truth or higher scores (indicating worse performance) when their predictions were far from the truth. Similar patterns in ranking and relative model performance were seen when stratifying ranks by pandemic phase (Figure S3).

**Figure 2:**
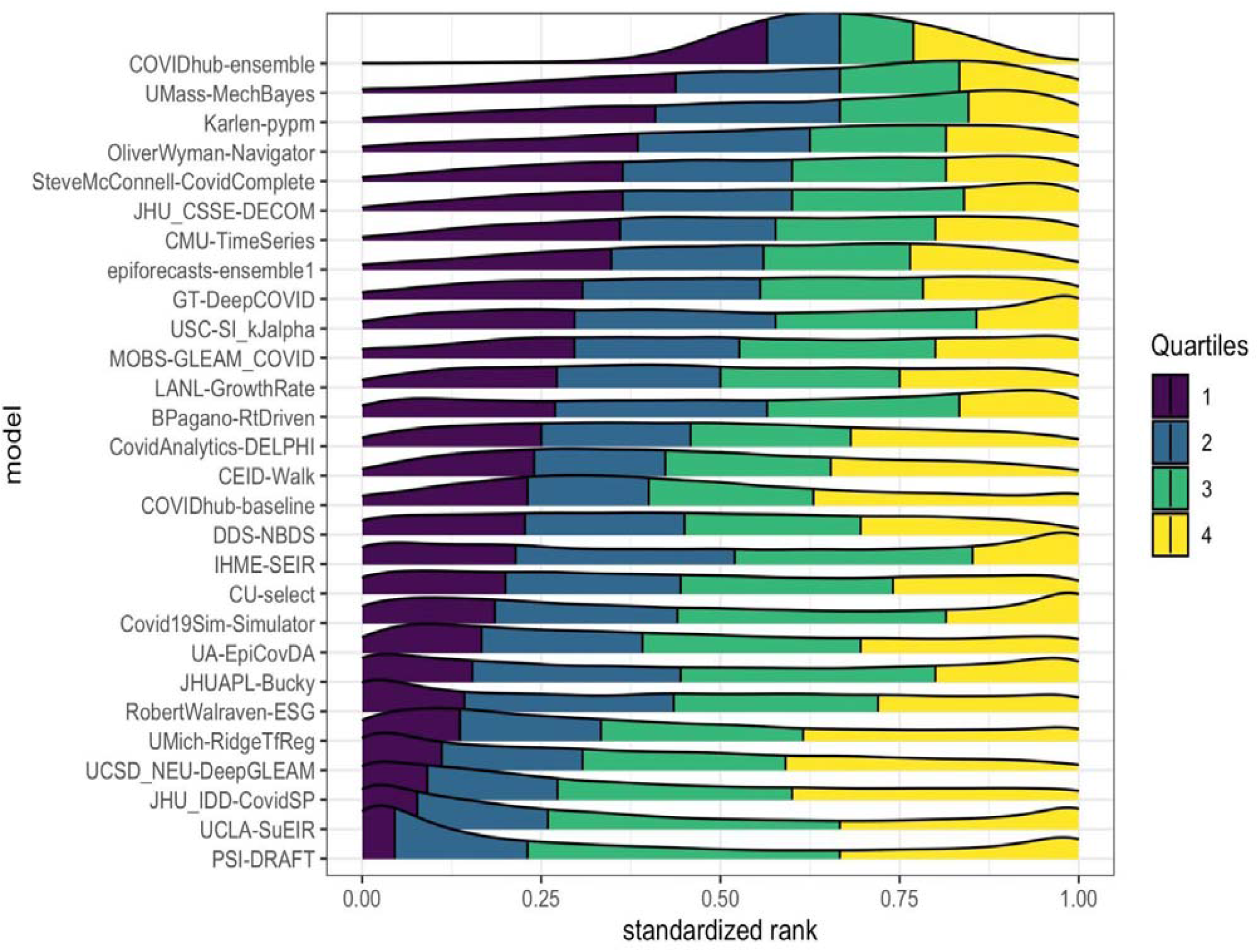
A comparison of each model’s distribution of standardized rank of weighted interval scores (WIS) for each location-target-week observation. A standardized rank of 1 indicates that the model had the best WIS for that particular location, target, and week and a value of 0 indicates it had the worst WIS. The density plots show interpolated distributions of the standardized ranks achieved by each model for every observation that model forecasted. The quartiles of each model’s distribution of standardized ranks are shown in different colors: yellow indicates the top quarter of the distribution and purple indicates the bottom quarter of the distribution. The models are ordered by the first quartile of the distribution, with models that rarely had a low rank near the top.

### Observations on accuracy in specific weeks

Forecasts from individual models showed variation in accuracy by forecast week and horizon (Figure 3). The COVIDhub-ensemble model showed better average WIS than both the baseline model and the average error of all models across the entire evaluation period, except for three weeks where the baseline had lower 1-week ahead error than the ensemble. The COVIDhub-ensemble 1-week ahead forecast for EW02-2021 yielded its highest average WIS across all weeks (average WIS = 72.7), and 9 out of 26 other models that submitted for the same locations outperformed it that week. The 4-week ahead COVIDhub-ensemble forecasts were worse in EW49-2020 than in any other week during the evaluation period (average WIS = 111.7), and 15 out of the 26 models outperformed the ensemble that week at a forecast horizon of 4 weeks.

**Figure 3:**
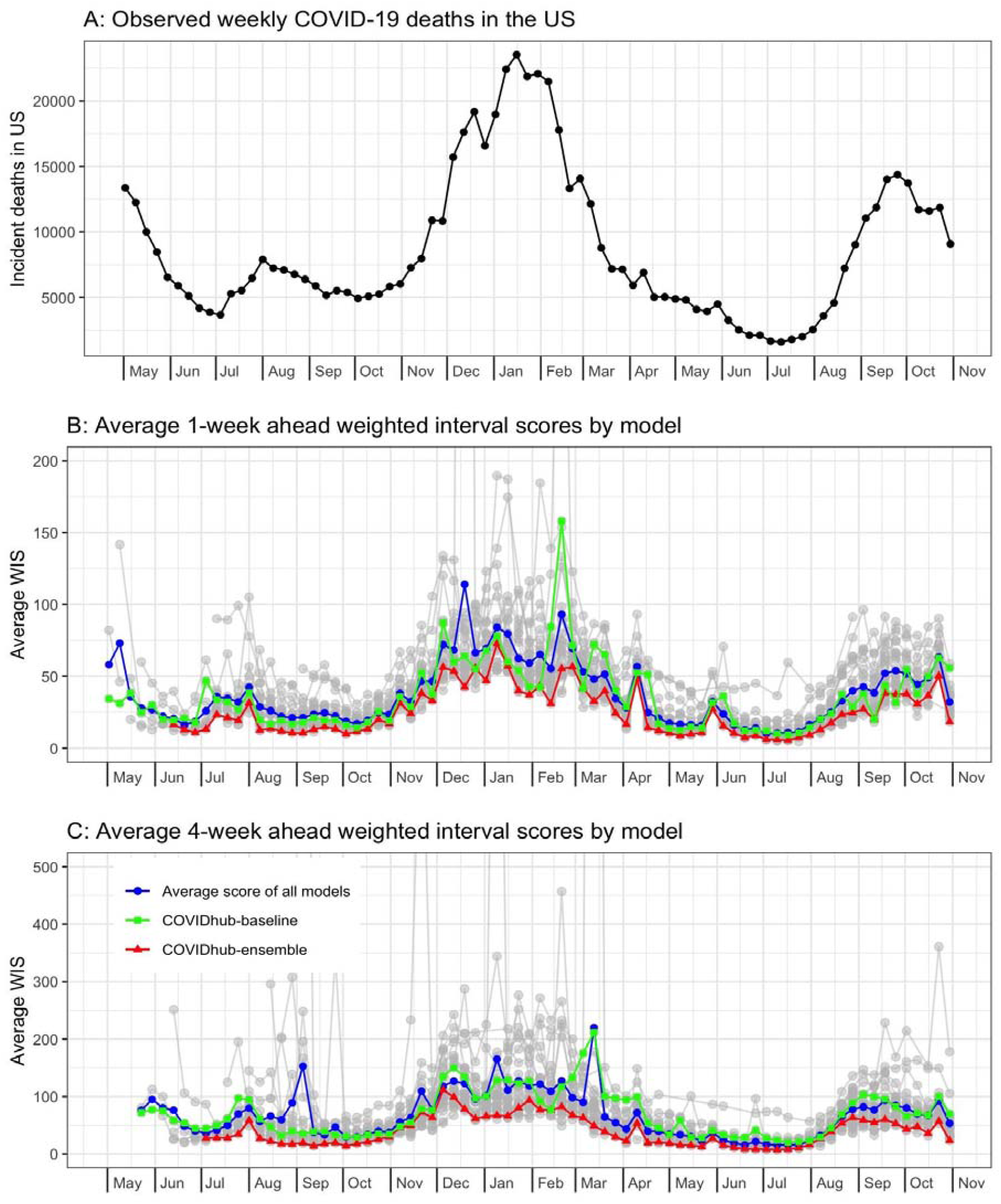
Average WIS by the target forecasted week for each model across all 50 states. Panel A shows the observed weekly COVID-19 deaths based on the CSSE reported data as of May 25, 2021. Panel B shows the average 1-week ahead WIS values per model (in grey). For all 21 weeks in which the ensemble model (red triangle) is present, this model has lower WIS values than the baseline model (green square) and the average score of all models (blue circle). The y-axes are truncated in panels B and C for readability of the majority of the data.

There was high variation among the individual models in their forecast accuracy during periods of increasing deaths and near peaks (i.e., forecast dates in July through early August of 2020, November through March, and August - October of 2021, Figure 3). High errors in the baseline model tended to be associated with large outliers in observed data for a particular week, e.g. times when a state reported a large backfill of deaths in the most recent week (SI Text). In general, other models did not show unusual errors in their forecasts originating from these anomalous data, suggesting that their approaches, including possible adjustments to recent observations, were robust to anomalies in how data were reported.

### Model performance in specific pandemic waves

In addition to evaluating performance in aggregate across the entire evaluation period and separate phases, we evaluated model performance during important moments during the pandemic. To assess the impact of rapidly changing trends on incident death forecasting accuracy, we ran an analysis restricted to specific locations and time periods that experienced high rates of change during four different waves of the pandemic (Figure 4):

**Figure 4:**
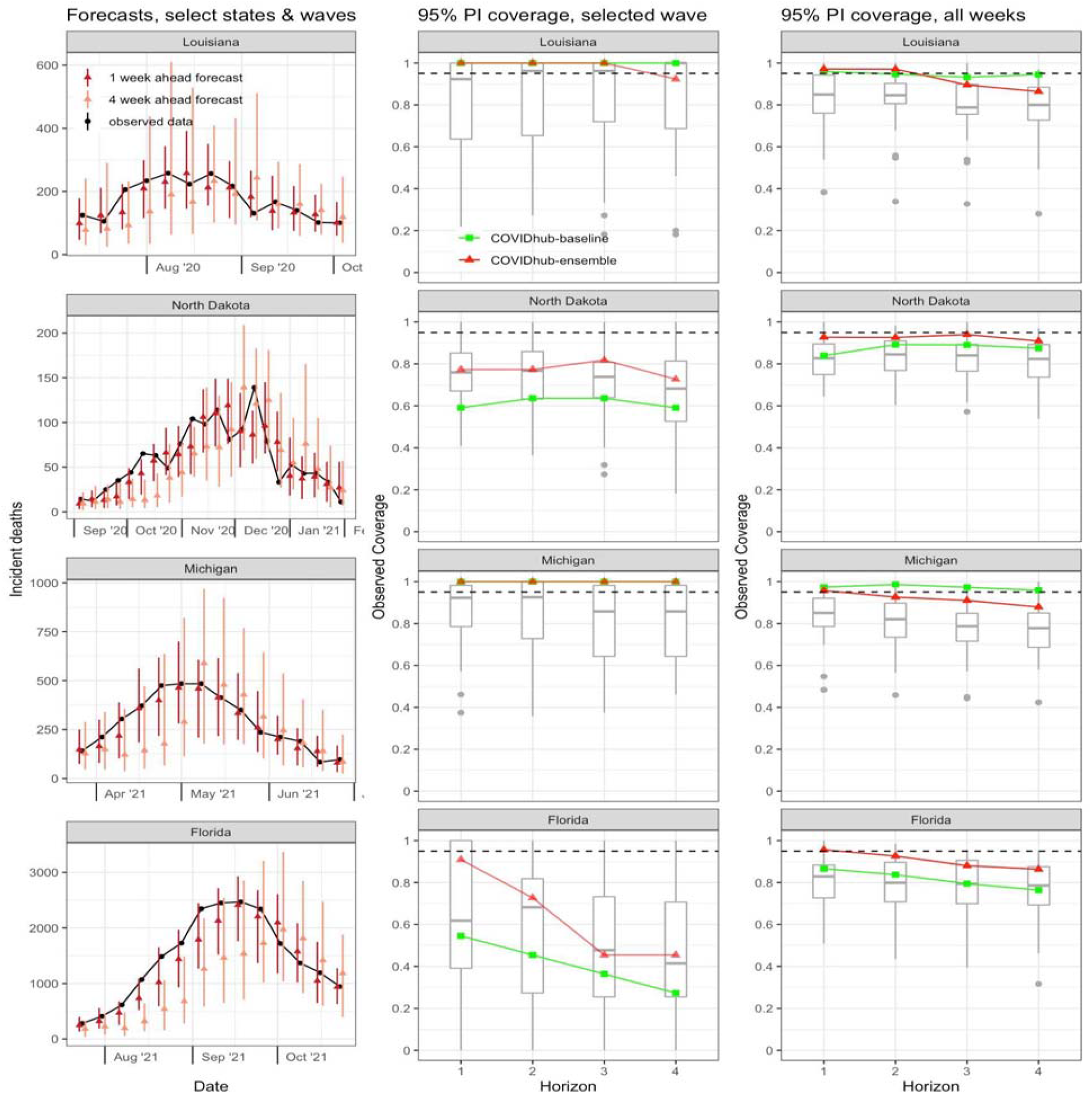
Forecasts for selected states and pandemic waves, with prediction interval coverage. The first column shows every 1 and 4 week ahead forecast with 95% prediction intervals (PIs) made by the ensemble during the selected evaluation period. The second and third columns of plots show evaluations of PIs, across 1 through 4 week horizons (x-axis). The red line with triangle points corresponds to the coverage rates of the COVIDhub-ensemble forecasts, green squares refer to the COVIDhub-baseline model. The boxplots represent the distribution of coverage rates from all component models. The second column evaluates only forecasts made for the dates shown in the first column. The third column evaluates forecasts across all weeks in the evaluation period. In the last two columns, the expected coverage rate (95%) is shown by the dashed line.

a. the summer 2020 waves in the south and southwest,
b. the late fall 2020 rise in deaths in the upper Midwest,
c. the wave driven by the Alpha SARS-CoV-2 variant in Michigan in in March/April 2021, and
d. the Delta variant wave in summer 2021 throughout most states in the US.

Forecast performance varied substantially in these examples. Models in general systematically underpredicted the mortality curve as trends were rising and overpredicted as trends were falling. In some of the selected waves (e.g., North Dakota and Florida), the ensemble forecast showed inappropriate levels of uncertainty, with the 95% prediction intervals covering the eventual observations less than 80% of the time. However, during other waves (e.g., Louisiana and Michigan) the ensemble forecast, while systematically biased first below and then above the eventually reported counts of deaths, did cover the observations at or above 95% of the time, although PIs were very wide. In general, lower-than expected coverage rates and bias were more pronounced at a 4-week horizon than a 1-week horizon. These four examples appeared to be representative of trends observed when looking across a larger number of waves (Dataset S2).

### Individual model forecast performance varies substantially by location

Forecasts from individual models showed large variation in accuracy by location when aggregated across all weeks and targets (Figure 5). Only the ensemble model showed superior accuracy when compared to baseline in all locations. Ensemble forecasts of incident deaths showed the largest relative accuracy improvements in New York, New Jersey, Indiana (relative WIS = 0.4), California, Massachusetts, and at the national level (relative WIS = 0.5), and the lowest relative accuracy in Vermont, Guam, and The Virgin Islands (relative WIS = 0.9). The COVIDhub-ensemble was the only model to outperform the baseline in every location when eligible in a specific pandemic phase (Figure S6).

**Figure 5:**
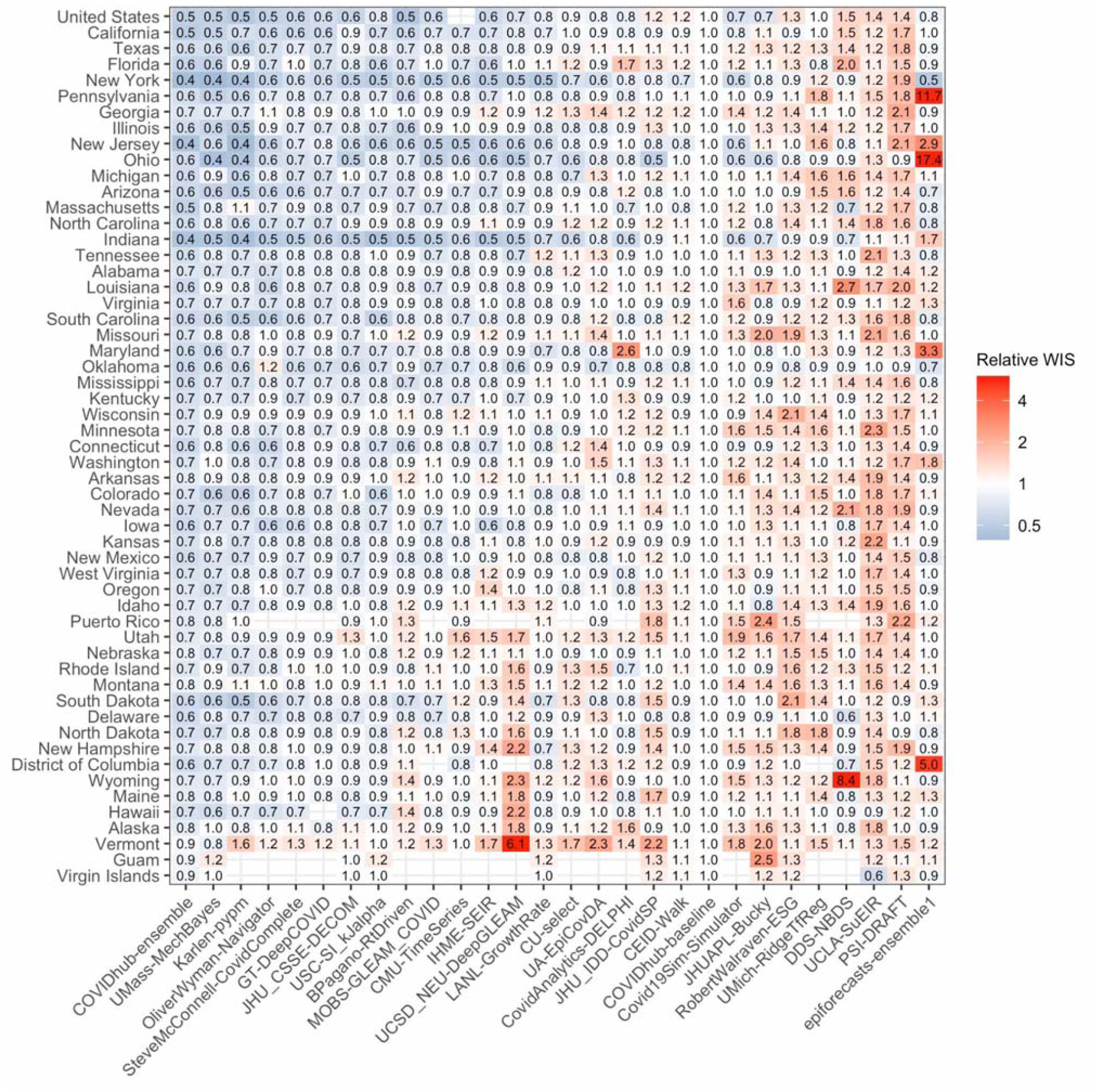
Relative WIS by location for each model across all horizons and submission weeks. The value in each box represents the relative WIS calculated from 1- to 4-week ahead targets available for a model at each location. Boxes are colored based on the relative WIS compared to the baseline model. Blue boxes represent teams that outperformed the baseline and red boxes represent teams that performed worse than the baseline. Locations are sorted by cumulative deaths as of the end of the evaluation period (October 30, 2021). Teams are listed on the horizontal axis in order from the lowest to highest relative WIS values (Table 1).

### Forecast performance degrades with increasing horizons

Averaging across all states and weeks in the evaluation period, forecasts from all models showed lower accuracy and higher variance at a forecast horizon of 4 weeks ahead compared to a horizon of 1 week ahead; however, models generally showed improved performance relative to the naive baseline model at larger horizons (Figure S4). Eleven models showed a lower average WIS (range: 24.9-34.3) than the baseline at a 1-week horizon (average WIS = 35.8). At a 4-week ahead horizon, 19 models had a lower average WIS (range: 39.9-65.2) than baseline (average WIS = 70.1). Across all models except one, the average WIS was higher than the median WIS, indicative of outlying forecasts impacting the mean value.

When averaging across locations and stratifying by phase of the pandemic, there was variation in the top performing models (Figure S5). Four models had a lower mean wis than baseline for both 1- and 4-week ahead targets in at least three out of four phases (COVIDhub-ensemble, GT-DeepCOVID, Karlen-pypm, UMass-MechBayes). Additionally, UMass-MechBayes, and COVIDhub-ensemble were the only models to appear in the top three models in three of the four phases analyzed (Figure S5). In contrast to average WIS, prediction interval coverage rates did not change substantially across the 1- to 4-week horizons for most models (Figure S2).

While many teams submitted only short-term (1 to 4 week ahead) forecasts, a smaller number of teams consistently submitted longer-term predictions with up to a 20-week horizon for all 50 states (Figure S8). Across all teams submitting forecasts for the 50 states, 4-week ahead forecasts had around 76% more error (based on relative WIS) than 1-week ahead forecasts, a relationship that was consistent across the entire evaluation period. Longer-term forecasts showed less accuracy on average than 1- and 4-week ahead forecasts. There were no clear overall differences in probabilistic model accuracy between 8- and 20-week horizons, although in early summer 2020, late spring 2021, and fall of 2021 average WIS at 8-week horizons were slightly lower than at longer horizons (Figure S8B). For the two teams who made 20-week ahead forecasts for all 50 states, average WIS was 2.9 to 4 times higher at a 20-week horizon than it was at a 1-week horizon. The increased WIS at longer prediction horizons for these models were due to larger dispersion (i.e. wider predictive distributions representing increased uncertainty) as well as larger penalties for underprediction and overprediction (Figure S9). The biggest increases in WIS were from increased penalties for underprediction, suggesting that the model forecasts did not accurately capture the possibility of increases in incidence at long horizons. Coverage rates for 95% prediction intervals tended to be stable or decline as the horizon increased (Figure S8C).

## Discussion

Given the highly visible role that forecasting has played in the response to the COVID-19 pandemic, it is critical that consumers of models, such as decision-makers, the general public, and modelers themselves, understand how reliable models are. This paper provides a comprehensive and comparative look at the probabilistic accuracy of different modeling approaches for forecasting COVID-19-related deaths during the COVID-19 pandemic in the US from April 2020 through October 2021. This work illustrates the tension between the desire for long term forecasts, which would be helpful for public health practitioners, and the decline in forecast accuracy at longer horizons that is shown by all forecasting methods.

As has been shown in prior epidemic forecasting projects, ensemble forecasts streamline and simplify the information provided to model consumers, and can provide a stable, accurate, and low-variance forecast (3, 13–15). The results presented here, which show high variation in accuracy between and within stand-alone models but more consistent accuracy from an ensemble forecast, support these prior results and confirm that an ensemble model can provide a reliable and comparatively accurate means of forecasting that exceeds the performance of most, if not all, of the models that contribute to it. The ensemble approach was the only model that (a) outperformed the baseline forecast in every location, (b) had better overall 4-week-ahead accuracy than the baseline forecast in every week, and (c) ranked in the top half of forecasts for more than 85% of the forecasts it made. Additionally, it achieved the best overall measures of point and probabilistic forecast accuracy for forecasting deaths. However, during key moments in the pandemic, while the ensemble outperformed many models, it often showed lower than desired accuracy (Figures 3 and 4).These results continue to strengthen the evidence base for synthesizing multiple models for public health decision support.

We summarize the key findings of the work as follows.

- The performance of all individual models forecasting COVID-19 mortality was highly variable, even for short-term targets (Figures 2 and 3). One source of variation was the data inputs. Further investigation is needed to determine in what settings additional data can yield measurable improvements in forecast accuracy or add valuable diversity to a collection of models that are being combined.
- A simple ensemble forecast that combined all submitted models each week was consistently the most accurate model when aggregating performance across forecast targets (Figure S4), weeks (Figure 3), or locations (Figure 5). Although rarely the “most accurate” model for individual predictions, the ensemble was consistently one of the top few models for any single prediction (Figure 2). For public health agencies concerned with using a model that shows dependably accurate performance, this is a desirable feature of a model.
- The high variation in ranks of models for each location-target-week suggests that all models, even those that are not as accurate on average, have observations for which they are the most accurate (Figure 2).
- The post hoc evaluation of models during forecasting waves in select states showed poor accuracy of the ensemble model’s point forecasts for 1- and 4-week ahead. During periods of increasing incident deaths the ensemble tended to underpredict, and during periods of decreasing incident deaths the ensemble tended to overpredict. Prediction interval coverage during these times varied (Figure 4).
- Forecast accuracy and calibration were substantially degraded as forecast horizons increased, largely due to underestimating the possibility of increases in incidence at long horizons (Figures S8 and S9).

It is critical that model performance is assessed both in aggregate (to assess models that showed the best overall performance) and in specific important moments during the pandemic. It is of public health interest to evaluate how well models are able to predict points at which the observed trends change. However, we note that a post-hoc evaluation that focuses only on times where a specific type of trend was observed raises conceptual challenges. Extreme turning points in the pandemic are relatively rare compared with the many weeks where trends continue or only slightly change from previous weeks. A post-hoc evaluation that focuses exclusively on these “change points” may reward models that may regularly predict extreme changes even when they do not occur at other times (20). Adapting proper scoring rules to weigh good performance in both kinds of situations is difficult.

Rigorous evaluation of forecast accuracy faces many limitations in practice. The large variation and correlation in forecast errors across targets, submission weeks, and locations) makes it difficult to create simple and rigorous comparisons of models. Forecast comparison is also challenging because teams have submitted forecasts for different lengths of time, different locations, and for different numbers of horizons (Figures S8 and S1). Some teams have also changed their models over time (Tables S1 and S2, Figure S1). To account for some of this variability, we implemented specific inclusion criteria. However, those criteria may exclude valuable approaches that were not applied to a large fraction of locations or weeks (see Methods).

Forecast performance may be affected by ground truth data and forecast target. Ground truth data are not static. They can be later revised as more data become available (Dataset S1). There are also instances where data are not revised but rather left with large peaks or dips due to reporting effects, especially around holidays. Different sources for ground truth data can also have substantial differences that impact model performance. Lastly, because this evaluation focuses on incident death forecasts, it cannot speak to model performance for incident cases or hospitalizations. Deaths may serve as a lagging indicator of COVID-19, thus making it more predictable than hospitalization and case targets (21).

While the Hub has provided many insights into what has and has not been predictable in the COVID-19 pandemic, it also has left many important questions unanswered. Due to the operational, real-time orientation of the project, the Hub has not collected data on experimental modeling studies where certain features can be included or left out to explicitly test what features of a model increase predictive accuracy. An observational study could be conducted with forecasts collected by the hub, but any such analysis would likely be confounded by other factors about how the model was built and validated. Other research in this area has shown small but measurable improvements in predictive accuracy by including other data streams available in real time (22). Continued research in this area is needed, especially to evaluate how behavioral, mobility, variant prevalence, or other data streams, might enhance predictive modeling.

Short-term forecasts of COVID-19 mortality have informed public health response and risk communication for the pandemic. The number of teams and forecasts contributing to the COVID-19 ensemble forecast model has exceeded forecasting activity for any prior epidemic or pandemic. However, these forecasts are only one component of a comprehensive public health data and modeling system needed to help inform outbreak response. Preparedness for future pandemics could be facilitated by creating template infrastructure and resources for arriving at and maintaining model submission formats. This project underscores the role that collaboration and active coordination between governmental public health agencies, academic modeling teams, and industry partners can play in developing modern modeling capabilities to support local, state, and federal response to outbreaks.

## Methods

### Surveillance data

Early in the COVID-19 pandemic, the Johns Hopkins Center for Systems Science and Engineering (CSSE) developed a publicly available data tracking system and dashboard that was widely used (23). CSSE collected daily data on cumulative reported deaths due to COVID-19 at the county, state, territorial, and national levels and made these data available in a standardized format beginning in March 2020. Incident deaths were inferred from this time-series as the difference in reported cumulative deaths on successive days. Throughout the real-time forecasting exercise described in this paper, the Forecast Hub stated that forecasts of deaths would be evaluated using the CSSE data as the ground truth and encouraged teams to train their models on CSSE data.

Like data from other public health systems, the CSSE data occasionally exhibited irregularities due to reporting anomalies. CSSE made attempts to redistribute large “backlogs” of data to previous dates in instances where the true dates of deaths, or dates when the deaths would have been reported, were known. However, in some cases, these anomalous observations were left in the final dataset (SI Text). All updates were made available in a public GitHub repository (https://github.com/CSSEGISandData/COVID-19/tree/master/csse_covid_19_data#data-modification-records). Weekly incidence values were defined and aggregated based on daily totals from Sunday through Saturday, according to the standard definition of epidemiological weeks (EW) used by the CDC (24).

### Forecast format

Research teams from around the world developed forecasting models and submitted their predictions to the COVID-19 Forecast Hub, a central repository that collected forecasts of the COVID-19 pandemic in the US beginning in April 2020. The Forecast Hub submission process has been described in detail elsewhere (25). Incident death forecasts, the focus of this evaluation, could be submitted with predictions for horizons of 1-20 weeks after the week in which a forecast was submitted.

A prediction for a given target (e.g., “1-week ahead incident death”) and location (e.g., “California”) was specified by one or both of a point forecast (a single number representing the prediction of the eventual outcome) and a probabilistic forecast. Probabilistic forecasts were represented by a set of 23 quantiles at probability levels 0.01, 0.025, 0.05, 0.10, 0.15, …, 0.95, 0.975, 0.99.

### Forecast model eligibility and evaluation period

To create a set of standardized comparisons between forecasts, only models that met specific inclusion criteria were included in the analysis. For the 79 weeks beginning in EW17-2020 and ending with EW42-2021, a model’s weekly submission was determined to be eligible for evaluation if the forecast

1. was designated as the “primary” forecast model from a team (groups who submitted multiple parameterizations of similar models were asked to designate prospectively a single model as their scored forecast);
2. contained predictions for at least 25 out of 51 focal locations (national level and states);
3. contained predictions for each of the 1-through 4-week ahead targets for incident deaths; and
4. contained a complete set of quantiles for all predictions.

A model was included in the evaluation if it had submitted an eligible forecast for at least 60% (n=47) of the submission weeks during the continuous 79 week period (Figure S1). Based on the eligibility criteria, we compared 28 models that had at least 47 eligible weeks during this time period.

### Aggregated forecast evaluation of pandemic phases

In a secondary analysis, forecasts were evaluated based on model submissions during four different phases of the pandemic. A model was eligible for inclusion in a given phase if it met the eligibility criteria listed above and had forecast submissions for at least 60% of the weeks during that phase. For the spring phase, models had to submit eligible forecasts for at least six out of 10 weeks starting EW16-2020 and ending EW26-2020. For summer eligibility, a model required submissions for at least 12 out of 20 submission weeks between EW27-2020 and EW46-2020. For winter eligibility, a model required submissions for at least 14 out of 23 submission weeks between EW47-2020 and EW16-2021. For delta phase eligibility, a model required submissions for at least 16 out of 26 submission weeks between EW17-2021 and EW42-2021. These phases were determined based on the waves of deaths at the national level during pandemic (Figure 1b). Each phase includes a period of increasing and decreasing incident deaths, although forecasts for the spring phase did not begin early enough to capture the increase in many locations.

Forecasts were scored using CSSE data available as of November 16, 2021. We did not evaluate forecasts on data first published in the 2 weeks prior to this date due to possible revisions to the data.

### Disaggregated forecast evaluation by pandemic wave

In a post-hoc secondary analysis, we evaluated forecasts made in selected locations during selected pandemic waves. We used the following criteria in selecting locations and waves to represent this analysis (Figure 4, Dataset S2).

- We selected states that had unusually severe waves or whose waves “led” the overall wave. Locations for which data for weekly deaths during the wave had been substantially revised after the initial report were excluded from consideration.
- We picked an initial date near the start of the first increase at the start of the wave and a last date at the end of the steep decline of the wave.

To compare the forecasts during the waves, we plotted 1 and 4 week ahead forecasts and calculated 95% prediction interval coverage rates of forecasts made for the given location both during the wave of interest over all weeks. Coverage rates were computed for models that were included in the overall analysis (see eligibility criteria above) and, for inclusion in the coverage calculations for each wave, the model additionally had to have made forecasts for at least 3 weeks in the selected wave.

### Forecast locations

Forecasts were submitted for 57 locations including all 50 states, 6 jurisdictions and territories (American Samoa, Guam, the Northern Mariana Islands, US Virgin Islands, Puerto Rico, and the District of Columbia), and a US national level forecast. Because American Samoa and the Northern Mariana Islands had no reported COVID-19 deaths and one reported COVID-19 death respectively during the evaluation period, we excluded these locations from our analysis.

In analyses where measures of forecast skill were aggregated across locations, we typically only included the 50 states in the analysis. Including these territories in raw score aggregations would favor models that had forecasted for these regions because models were often accurate in predicting low or zero deaths each week, thereby reducing their average error. The national level forecasts were not included in the aggregated scores because the large magnitude of scores at the national level strongly influences the averages. However, in analyses where scores were stratified by location, we included forecasts for all US states, including territories and the national level.

This evaluation used the CSSE COVID-19 surveillance data as ground truth when assessing forecast performance. We did not score observations when ground-truth data showed negative values for weekly incident deaths (due to changes in reporting practices from state/local health agencies, e.g., removing “probable” COVID-19 deaths from cumulative counts). This occurred 11 times.

### Forecast models

For the primary evaluation, we compared 28 models that submitted eligible forecasts for at least 47 of the 79 weeks considered in the overall model eligibility period (Figure 1). Teams that submitted to the COVID-19 Forecast Hub used a wide variety of modeling approaches and input data (Tables S1 and S2). Two of the evaluated models are from the COVID-19 Forecast Hub itself: a baseline model and an ensemble model.

The COVIDhub-baseline model was designed to be a neutral model to provide a simple reference point of comparison for all models. This baseline model forecasted a predictive median incidence equal to the number of reported deaths in the most recent week (*y*_*t*_), with uncertainty around the median based on changes in weekly incidence that were observed in the past of the time series (details in SI Text).

The COVIDhub-ensemble model combined forecasts from all models that submitted a full set of 23 quantiles for 1-through 4-week ahead forecasts for incident deaths. The ensemble for incident weekly deaths was first submitted in the week ending June 06, 2020 (EW23). For submission from EW23 through EW29 (week ending July 18, 2020), the ensemble took an equally weighted average of forecasts from all models at each quantile level. For submissions starting in EW30 (week ending July 25, 2020), the ensemble computed the median across forecasts from all models at each quantile level (26). We evaluated more complex ensemble methods, and while they did show modest improvements in accuracy, they also displayed undesirable increases in variability in performance during this evaluation period (27, 28).

### Forecast submission timing

Of the 3,555 forecast submissions we included in the evaluation, 230 (6%) were either originally submitted or updated more than 24 hours after the submission deadline. In all of these situations, modeling teams attested (via annotation on the public data repository) to the fact that they were correcting inadvertent errors in the code that produced the forecast, and that the forecast used as input only data that would have been available before the original submission due date. In these limited instances, we evaluated the most recently submitted forecasts.

### Evaluation methodology

We evaluated aggregate forecast skill using a range of scores of forecast skill that assessed both point and probabilistic accuracy. These scores were aggregated over time and locations for near-term forecasts (4 weeks or less into the future) and, in a single analysis, for longer-term projections (5-20 weeks into the future).

Point forecast error was assessed using the mean absolute error (MAE), defined for a set of observations *y*_1*:N*_ and each model’s designated point predictions *ŷ*_1*:N*_ as 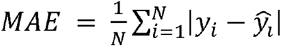

To assess probabilistic forecast accuracy, we used two scores that are easily computable from the quantile representation for forecasts described above. Briefly, the weighted interval score (WIS) is a proper score that combines a set of interval scores for probabilistic forecasts that provide quantiles of the predictive forecast distribution (19). Proper scores promote “honest” forecasting by not providing forecasters with incentives to report forecasts that differ from their true beliefs about the future (29). We also evaluated the prediction interval coverage, the proportion of times a prediction interval of a certain level covered the observed value, to assess the degree to which forecasts accurately characterized uncertainty about future observations. Details of the calculation of the WIS score and prediction interval coverage are provided in SI Text.

### Forecast comparisons

Comparative evaluation of the considered models 1, …, *M* is hampered by the fact that not all of them provide forecasts for the same set of locations and time points. To adjust for the level of difficulty of each model’s set of forecasts, we computed (a) a standardized rank between 0 and 1 for every forecasted observation relative to other models that made the same forecast, and (b) an adjusted relative WIS and MAE.

To compute the WIS standardized rank score for model *m* and observation *i*(*sr*_*m,i*_), we computed the number of models that forecasted that observation (*n*_*i*_) and the rank of model *m* among those *n*_*i*_ models (*r*_*m,i*_). The model with the best (i.e., lowest) WIS received a rank of 1 and the worst received a rank of *n*_*i*_. The standardized rank then rescaled the ranks to between 0 and 1, where 0 corresponded to the worst rank and 1 to the best, (30–32) as follows:

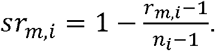

This metric is not dependent on the scale of the observed data. If all models were equally accurate, distributions of standardized ranks would be approximately uniform.

A procedure to compute a measure of relative WIS, which evaluates the aggregate performance of one model against the baseline model is described in SI Text. To adjust for the relative difficulty of beating the baseline model on the covered set of forecast targets, the chosen measure also takes into account the performance of all other available models. The same procedure was used to compute a relative MAE.

## Data Availability

All data and code referred to in the manuscript are publicly available.

https://github.com/reichlab/covid19-forecast-hub/

https://github.com/reichlab/covidEnsembles

https://zoltardata.com/project/44

https://github.com/reichlab/covid19-forecast-evals/blob/main/figures/data_revisions.pdf

## Data and code availability

The forecasts from models used in this paper are available from the COVID-19 Forecast Hub GitHub repository (https://github.com/reichlab/covid19-forecast-hub) (4) and the Zoltar forecast archive (https://zoltardata.com/project/44)(33). The code used to generate all figures and tables in the manuscript is available in a public repository (https://github.com/reichlab/covid19-forecast-evals). All analyses were conducted using the R language for statistical computing (v 4.0.2) (34).

We followed the EPIFORGE 2020 guidelines for reporting results from epidemiological forecasting studies (Table S5) (35).

## Funding

For teams that reported receiving funding for their work, we report the sources and disclosures below. The content is solely the responsibility of the authors and does not necessarily represent the official views of any of the funding agencies.

*<u>CMU-TimeSeries</u>*: CDC Center of Excellence, gifts from Google and Facebook.

*<u>CU-select</u>*: NSF DMS-2027369 and a gift from the Morris-Singer Foundation.

*<u>COVIDhub</u>*: US CDC (1U01IP001122); NIGMS (R35GM119582). Helmholtz Foundation (SIMCARD Information & Data Science Pilot Project). Klaus Tschira Foundation.

*<u>Columbia_UNC-SurvCon:</u>* GM124104

*<u>DDS-NBDS</u>*: NSF III-1812699.

*<u>EPIFORECASTS-ENSEMBLE1</u>*: Wellcome Trust (210758/Z/18/Z)

*<u>GT_CHHS-COVID19</u>*: William W. George Endowment, Virginia C. and Joseph C. Mello Endowments, NSF DGE-1650044, NSF MRI 1828187, CDC and CSTE NU38OT000297, PACE at GATech. Andrea Laliberte, Joseph C. Mello, Richard “Rick” E. & Charlene Zalesky, and Claudia & Paul Raines.

*<u>GT-DeepCOVID</u>*: CDC MInD-Healthcare U01CK000531-Supplement. NSF (Expeditions CCF-1918770, CAREER IIS-2028586, RAPID IIS-2027862, Medium IIS-1955883, NRT DGE-1545362), CDC MInD program, ORNL and funds/computing resources from Georgia Tech and GTRI.

*<u>IHME</u>*: The Bill & Melinda Gates Foundation; the state of Washington and NSF (FAIN: 2031096).

*<u>IowaStateLW-STEM</u>*: Iowa State University Plant Sciences Institute Scholars Program, NSF DMS-1916204, NSF CCF-1934884, Laurence H. Baker Center for Bioinformatics and Biological Statistics.

*<u>JHU CSSE</u>*: NSF RAPID (2108526, 2028604)

*<u>JHU_IDD-CovidSP</u>*: State of California, US HHS, US DHS, US Office of Foreign Disaster Assistance, Johns Hopkins Health System, Office of the Dean JHBSPH, Johns Hopkins University Modeling and Policy Hub, CDC (5U01CK000538-03), University of Utah Immunology, Inflammation, & Infectious Disease Initiative (26798 Seed Grant).

*<u>LANL-GrowthRate</u>*: LANL LDRD 20200700ER.

*<u>MOBS-GLEAM_COVID</u>*: COVID Supplement CDC-HHS-6U01IP001137-01; CSTE Cooperative Agreement no. NU38OT000297.

*<u>NotreDame-mobility</u>*<u>and</u> *<u>NotreDame-FRED</u>*: NSF RAPID DEB 2027718

*<u>PSI-DRAFT</u>*: NSF RAPID Grant # 2031536.

*<u>UA-EpiCovDA</u>*: NSF RAPID DMS 2028401.

*<u>UCSB-ACTS</u>*: NSF RAPID IIS 2029626.

*<u>UCSD-NEU</u>*: Google Faculty Award, DARPA W31P4Q-21-C-0014, COVID Supplement CDC-HHS-6U01IP001137-01.

*<u>UMass-MechBayes</u>*: NIGMS R35GM119582, NSF 1749854.

*<u>UMich-RidgeTfReg</u>*: UMich Physics Department and Office of Research.

