## Supplemental Information for "Evaluation of individual and ensemble probabilistic forecasts of COVID-19 mortality in the US"

Cramer EY, Ray EL, et al.

Nicholas G Reich


**This PDF file includes:**

Supplementary text
Figures S1 to S9
Tables S1 to S5
Legends for Datasets S1 to S2
SI References

**Other supplementary materials for this manuscript include the following:**

Datasets S1 to S2

**Supplementary Information Text**

**Methods for calculating WIS and prediction interval coverage.** Given quantiles of a forecast distribution $F$, an observation $y$ and an uncertainty level $\alpha$, a single interval score is defined as

$$IS_{\alpha}(F, y) = (u - l) + \frac{2}{\alpha} \cdot (l - y) \cdot 1(y < l) + \frac{2}{\alpha} \cdot (y - u) \cdot 1(y > u)$$

where $1(\cdot)$ is the indicator function and $l$ and $u$ are the $\frac{\alpha}{2}$ and $1 - \frac{\alpha}{2}$ quantiles of $F$ (i.e., the lower and upper end of a central 1 - $\alpha$ prediction interval). Given a set of central prediction intervals, a weighted sum of interval scores can be computed to summarize accuracy across the entire predictive distribution. We define the WIS as a particular linear combination of $K$ interval scores, as

$$WIS_{\alpha_{0:K}}(F, y) = \frac{1}{K+1/2} \cdot \left( w_0 \cdot |y - m| + \sum_{k=1}^{K} w_k \cdot IS_{\alpha_k}(F, y) \right)$$

where $w_k = \frac{\alpha_k}{2}$ for $k = 1, \ldots, K$ and $w_0 = 1/2$. In our setting, we used $K = 11$ interval scores, for $\alpha$ =0.02, 0.05, 0.1, 0.2, …, 0.9.

This particular choice of weights for WIS is equivalent to the pinball loss used in quantile regression and has been shown to approximate the commonly used continuous ranked probability score (CRPS) (1). As such, it can be viewed as a distributional generalization of the absolute error, with smaller values of WIS corresponding to forecasts that are more consistent with the observed data (1, 2). WIS can be interpreted as a measure of how close the entire distribution is to the observation, in units on the scale of the observed data. We note that some alternative scores that are commonly used such as CRPS and the logarithmic score cannot be exactly calculated if only a set of quantiles of the predictive distribution are available.

An individual interval score for a single prediction and uncertainty level can be broken into three additive components, in order as they appear in the $IS$ equation above: dispersion, underprediction and overprediction. These components --dispersion, underprediction and overprediction as they appear respectively in the $IS$ equation above-- represent contributions to the score. As an example, say a 50% prediction interval ($\alpha = 0.5$) is (40, 60) and the true observation is 30. The $IS_{\alpha=0.5}(\{40, 60\}, 30) = 20 + 40 + 0 = 60$, where the dispersion is 20, the penalty for underprediction is 40, and there is no penalty for overprediction. Similarly, the WIS, which is computed as a weighted sum of interval scores across all available uncertainty levels, can similarly be split into contributions from each of these components. These then can be used to summarize the average performance of a model in terms of the width of its intervals and the average penalties it receives for intervals missing below or above the observation.

We note that average WIS scores are often driven by large errors in locations with high observed values. This approach assumes that a prediction of 1000 deaths when the truth is 500 should be seen as a more serious model failure than a prediction of 10 deaths when the truth is 5.

We also evaluated prediction interval coverage, the proportion of times a prediction interval of a certain level covered the observed value, to assess the degree to which forecasts accurately characterized uncertainty about future observations. While prediction interval coverage is not a proper score and only assesses one feature of a full predictive distribution, it does provide a clear and interpretable measure of forecast calibration. We compute prediction interval coverage for a set of observations ($y_i$, $i = 1, \ldots, N$) and prediction interval bounds with an uncertainty level $1 - \alpha$, $(l_{\alpha,i}, u_{\alpha,i})$, $i = 1, \ldots, N$ as

$$\text{prediction interval coverage} = \frac{1}{N} \sum_{i=1}^{N} 1(l_{\alpha,i} \le y_i \le u_{\alpha,i}).$$

We quote from Bracher et al (2021) (1) to illustrate the public health relevance of computing average error without population standardization:

> "The absolute error, when averaged across time and space, is dominated by forecasts from larger states and weeks with high activity (this also holds true for the CRPS and WIS). One may thus be tempted to use a relative measure of error instead, such as the mean absolute percentage error (MAPE). We argue, however, that emphasizing forecasts of targets with higher expected values is meaningful. For instance, there should be a larger penalty for forecasting 200 deaths if 400 are eventually observed than for forecasting 2 deaths if 4 are observed. Relative measures like the MAPE would treat both the same. Moreover, the MAPE does not encourage reporting predictive medians nor means, but rather obscure and difficult to interpret types of point forecasts [14,32]. It should therefore be used with caution."

**Methods for calculating relative WIS and relative MAE.** For each pair of models $m$ and $m'$, we computed the pairwise relative WIS skill

$$\theta_{mm'} = \frac{average\ WIS\ of\ model\ m}{average\ WIS\ of\ model\ m'}$$

based on the available overlap of forecast targets. Subsequently, we computed for each model the geometric mean of the results achieved in the different pairwise comparisons, denoted by

$$\theta_m = \left(\prod_{m'=1}^{M} \theta_{mm'}\right)^{1/M}.$$

Then, $\theta_m$ is a measure of the relative skill of model $m$ with respect to the set of all other models 1, …, $M$ , including the baseline. The central assumption here is that performing well relative to individual models 1, …, $M$ is similarly difficult for each week and location so that no model can gain an advantage by focusing on just some of them. As is, $\theta_m$ is a comparison to a hypothetical "average" model. Because we consider a comparison to the baseline model more straightforward to interpret, we rescaled $\theta_m$ and reported

$$\theta^*_m = \frac{\theta_m}{\theta_B},$$

where $\theta_B$ is the geometric mean of the results achieved by the baseline model in pairwise comparisons to all other models. The quantity $\theta^*_m$ then describes the relative performance of model $m$, adjusted for the difficulty of the forecasts model $m$ made, and scaled so the baseline model has a relative performance of 1. For simplicity, we refer to $\theta^*_m$ as the "relative WIS" or "relative MAE" throughout the manuscript. A value of $0 < \theta^*_m < 1$ means that model $m$ is better than the baseline, a value of $\theta^*_m > 1$ means that the baseline is better.

**Methods for creating baseline forecast.** The COVIDhub-baseline model was designed to be a neutral model to provide a simple reference point of comparison for all models. This baseline model forecasted a predictive median incidence equal to the number of reported deaths in the most recent week ($y_t$), with uncertainty around the median based on changes in weekly incidence that were observed in the past of the time series. This predictive distribution was created by collecting, for a particular location, the first differences and their negatives from the previously observed time series (i.e., $y_t - y_{t-1}$ and $-(y_t - y_{t-1})$ for all past times $t$). To obtain a smoother distribution of values to sample, we formed a distribution of possible differences based on a piecewise linear approximation to the empirical cumulative distribution function of the observed differences. We then obtained a Monte Carlo approximation of the distribution for incident deaths at forecast horizon *h* by independently sampling 100,000 changes in incidence at each week 1, 2, …, *h*, and adding sequences of *h* differences to the most recent observed incident deaths. Quantiles are reported for each horizon, with the median forced to be equal to the last observed value (to adjust for any noise introduced from the sampling process) and the distribution truncated so that it has no negative values.

**Outlying and anomalous observations.** We identified two main types of anomalies in the CSSE data: data revisions and outliers. We defined "data revisions" as observations that, after first being reported, were later substantially revised to a new value. A substantial revision was defined as one where (a) the absolute value of the observed difference between the original and updated observation was greater than 20, and (b) the relative difference was greater than or equal to 50%. We defined "outliers" as points that lay substantially far away from the reported data in nearby weeks based on review from two data experts. The goal was to identify observations that models should not be expected to predict accurately, either because the input data at a given time was not reliable due to later revisions or because the target data was not evaluable due to it being a substantial outlier. Supplemental analyses on forecasts between EW17-2020 through EW16-2021 showed that excluding revised or outlying observations did not substantially change the ordering of the models or the overall conclusions of the analysis (SI Table 4).

The extent to which initially reported observations were revised, in some cases multiple times, is documented in SI Dataset 1. When stratified by phase, it is more apparent that the quality of data impacted the relative performance of models in specific locations. For instance, in the winter 2020/2021 in Ohio, there were large data revisions (SI Dataset 1), which led to a more inaccurate baseline model. Therefore, nearly every model outperformed the baseline during this period. Additional locations in which large data revisions may have impacted baseline accuracy were observed in New York, New Jersey, and Indiana.

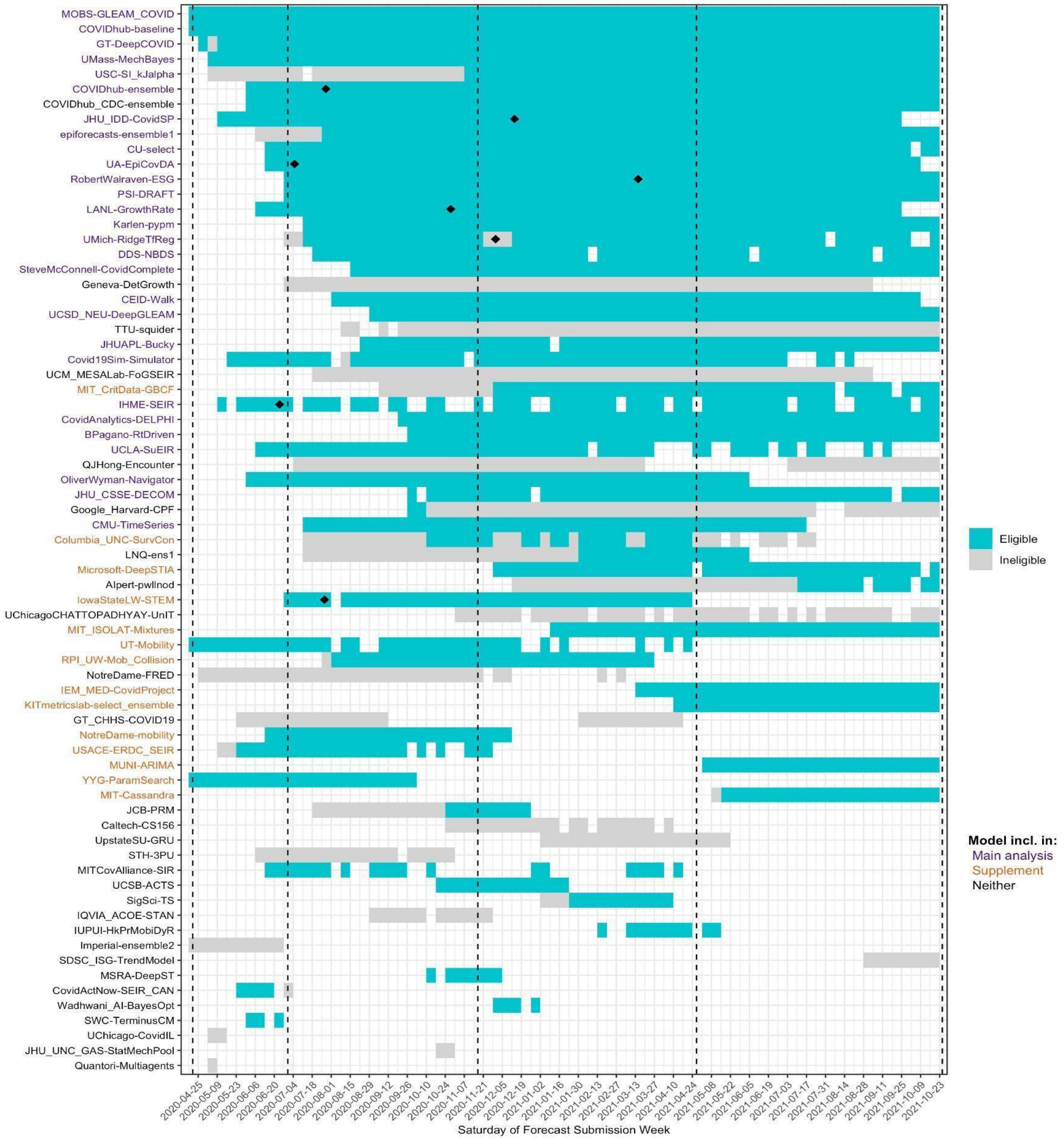


**Fig. S1.** Models contributed incident mortality forecasts to the COVID-19 Forecast Hub and were evaluated for eligibility for the analysis in this manuscript. Each cell represents the weekly submission from a particular model (row) in a particular week (column). Forecasts that were determined to be an eligible submission, based on forecasting for at least 25 locations and all of the 1 - 4 week horizons and submitting all quantiles, are highlighted in light blue. Submissions that are not eligible are shown in grey. Model names in purple indicate the teams included in the overall evaluation in the main text. Model names in orange indicate teams that were only included in a phase-specific evaluation included in the supplemental information. Models in black were not evaluated individually at any point. Vertical dashed lines demarcate the "phases" evaluated separately in the supplement. Black diamonds indicate the time points at which models were altered (Supplemental Table 1 footnotes).

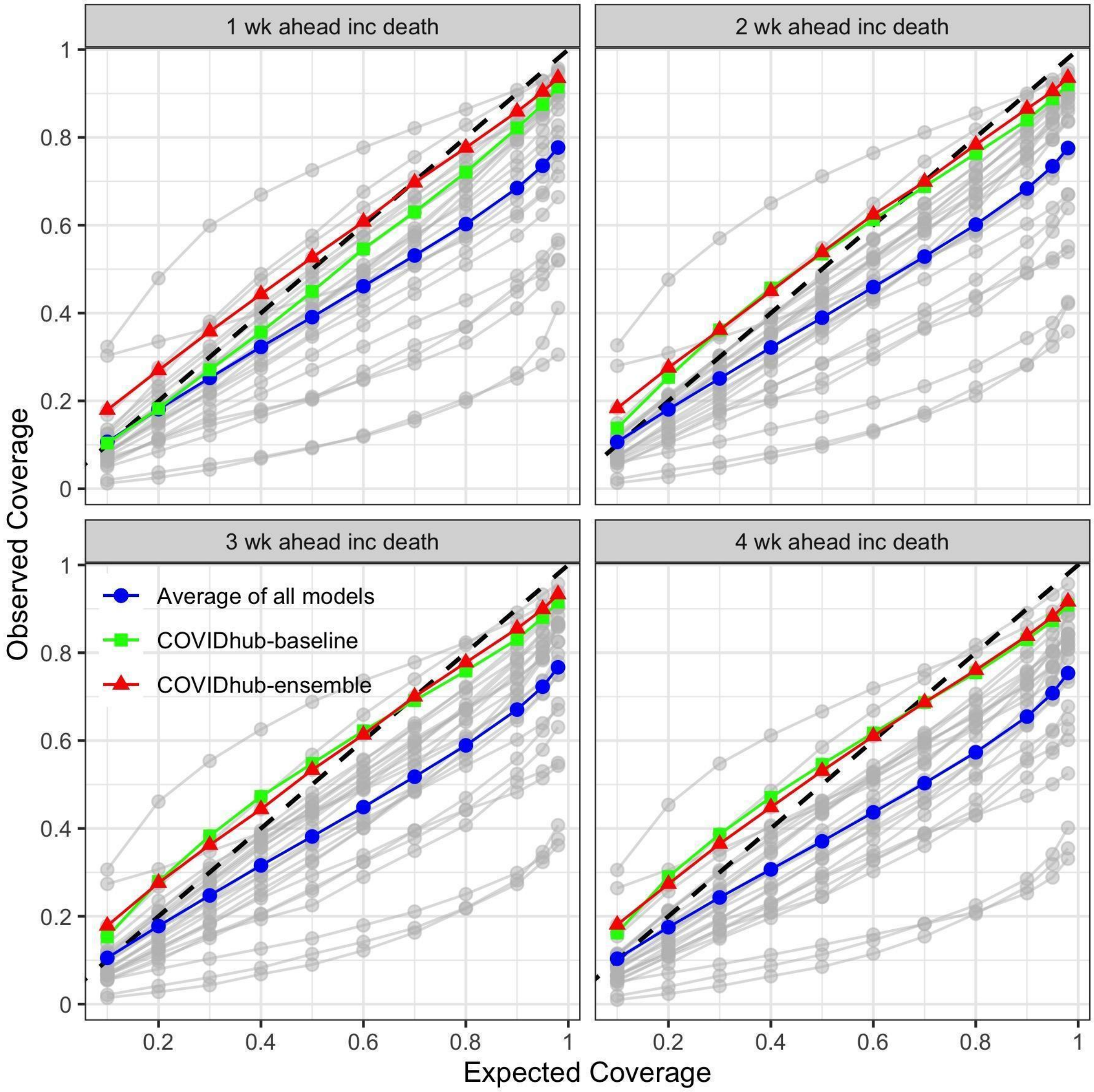


**Fig. S2.** Prediction interval coverage for all submitted forecasts with horizons of 1 through 4 weeks, aggregated across submission date, location, and week. Forecasts for any available forecasted location (nation, state, or territory) were included in this analysis. Points indicate PI coverage rates at nominal PI levels of 10%, 20%, 30%, ... 90%, 95%, and 98%. If a model is well calibrated across all PIs, the values should be close to the dashed black line, representing the expected PI coverage. As seen in each panel, few models (grey) have an observed coverage rate at or above the expected coverage rate. When averaged across all models (blue circle) the PI coverage falls below the expected coverage at every level at every horizon. The ensemble (red triangle) is better calibrated than the baseline model (green square) and the model average across nearly all PI levels for 1-week ahead and more than half of the levels for 4-week ahead horizons.

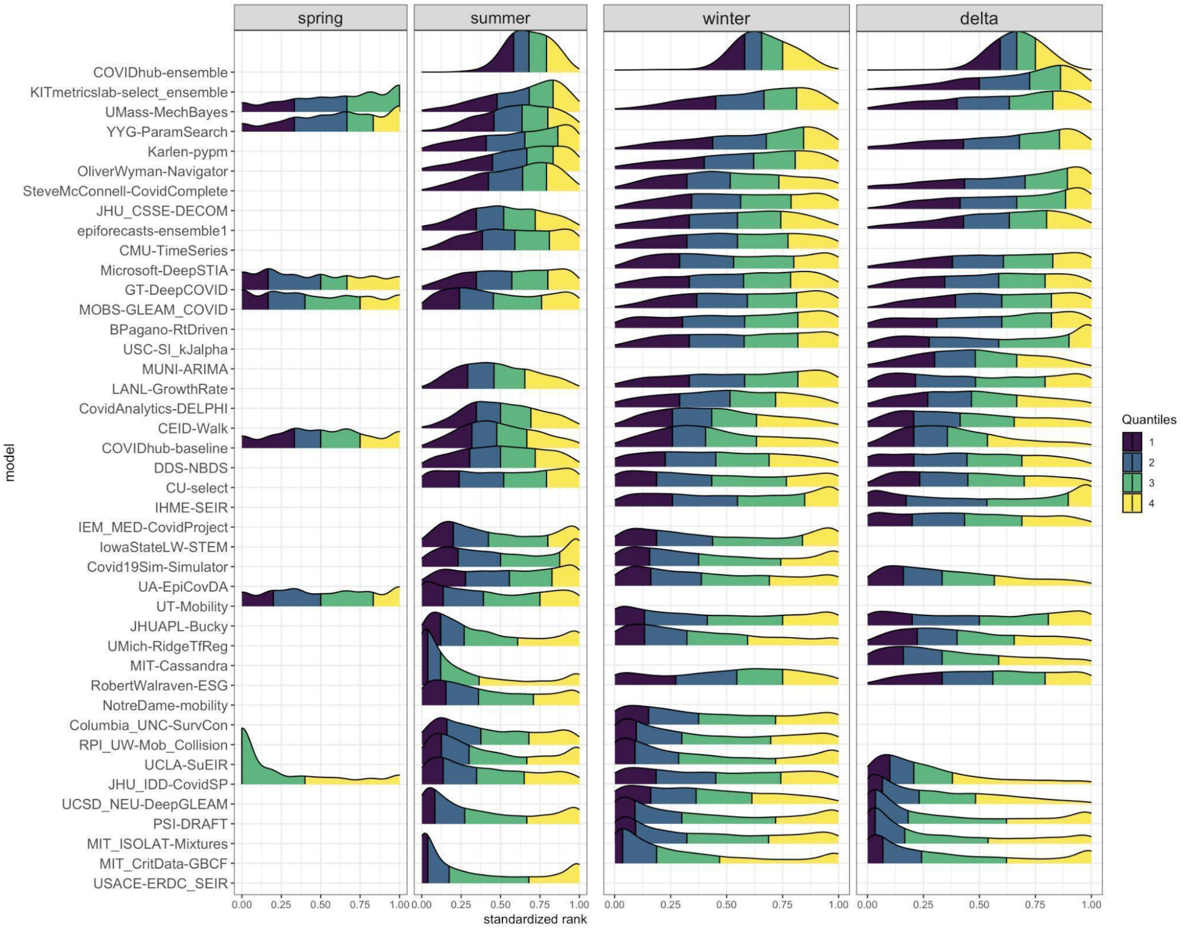


**Fig. S3.** A comparison of each model's distribution of standardized rank of weighted interval scores (WIS) for each location-target-week observation stratified by three phases of the pandemic. A standardized rank of 1 indicates that the model had the best WIS for that particular location, target, and week and a value of 0 indicates it had the worst WIS. The density plots show smoothly interpolated distributions of the standardized ranks achieved by each model for every observation that model forecasted. The quartiles of each model's distribution of standardized ranks are shown in different colors: yellow indicates the top quarter of the distribution and purple indicates the region containing the bottom quarter of the distribution. The models are ordered by the overall first quartile of the distribution with models that rarely had a low rank near the top. Observations in this figure included predictions for the national level, all 50 states, and 5 US territories. If models were equally accurate, all distributions would be approximately uniform.

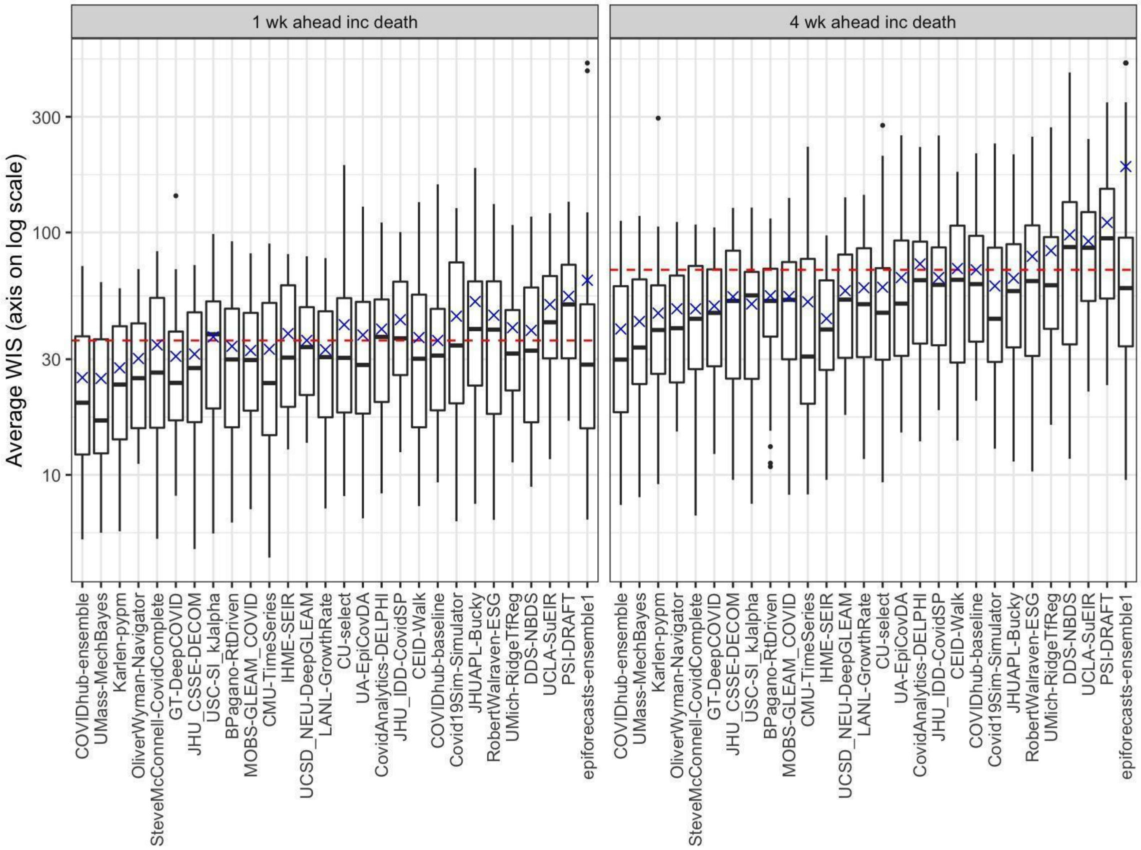


**Fig. S4.** Boxplot distributions of average weighted interval score (WIS, y-axis on log scale) for each week and model across all 50 states. The two panels represent 1 and 4 week ahead forecast horizons. The boxplots summarize the distribution of average WIS values for each week, averaging across all available locations and 1 - 4 week ahead horizons for each model. The “x” marks indicate the average WIS for each model. Models are ordered along the x-axis by their relative WIS (Table 1). The horizontal dashed lines indicate the average baseline WIS for each horizon.

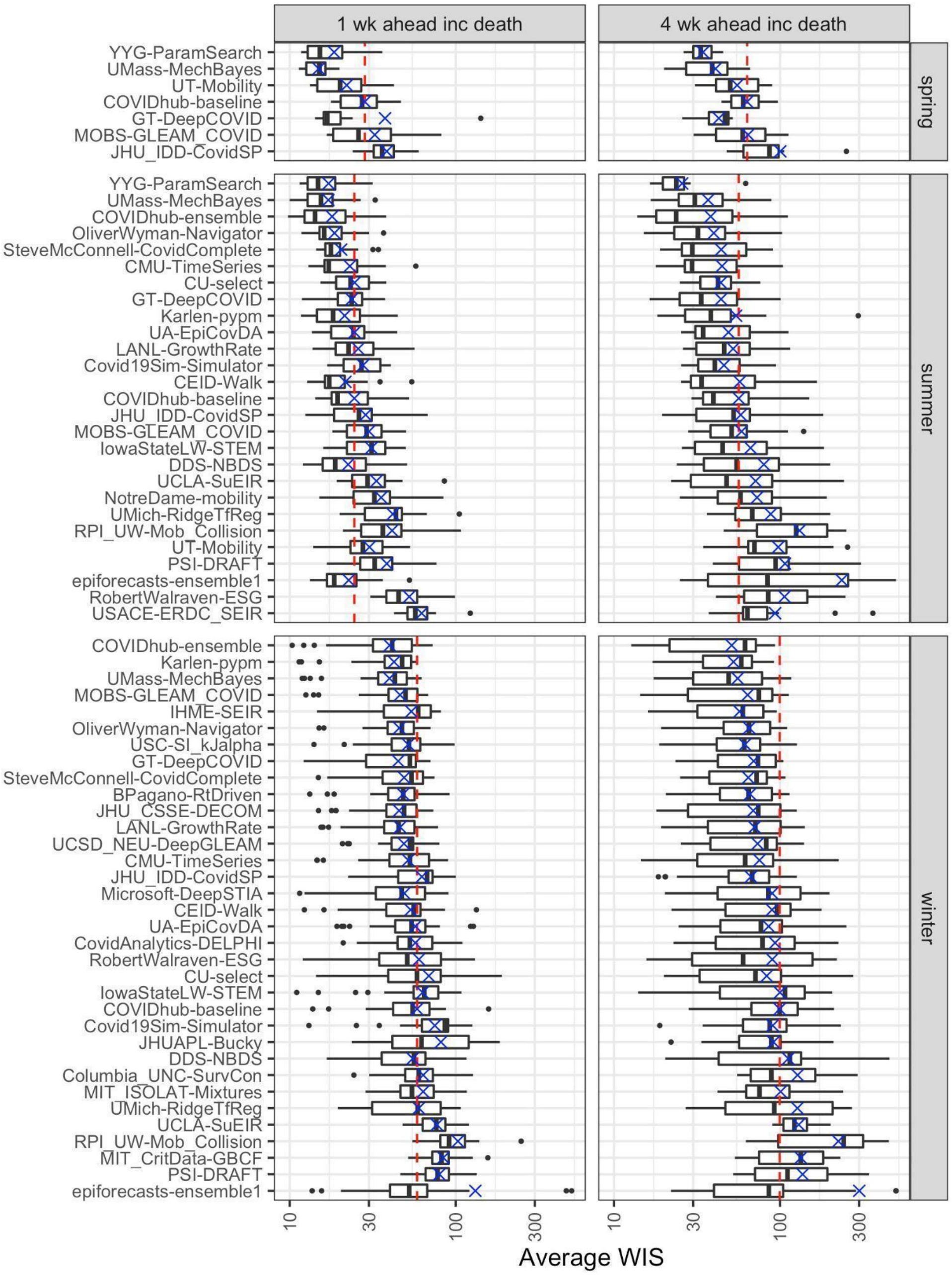

1 wk ahead inc death
4 wk ahead inc death
spring
summer
winter
YYG-ParamSearch
UMass-MechBayes
UT-Mobility
COVIDhub-baseline
GT-DeepCOVID
MOBS-GLEAM_COVID
JHU_IDD-CovidSP
COVIDhub-ensemble
OliverWyman-Navigator
SteveMcConnell-CovidComplete
CMU-TimeSeries
CU-select
Karlen-pypm
UA-EpiCovDA
LANL-GrowthRate
Covid19Sim-Simulator
CEID-Walk
IowaStateLW-STEM
DDS-NBDS
UCLA-SuEIR
NotreDame-mobility
UMich-RidgeTfReg
RPI_UW-Mob_Collision
PSI-DRAFT
epiforecasts-ensemble1
RobertWalraven-ESG
USACE-ERDC_SEIR
IHME-SEIR
USC-SI_kJalpha
BPagano-RtDriven
JHU_CSSE-DECOM
UCSD_NEU-DeepGLEAM
Microsoft-DeepSTIA
CovidAnalytics-DELPHI
JHUAPL-Bucky
Columbia_UNC-SurvCon
MIT_ISOLAT-Mixtures
MIT_CritData-GBCF
10
30
100
300
Average WIS

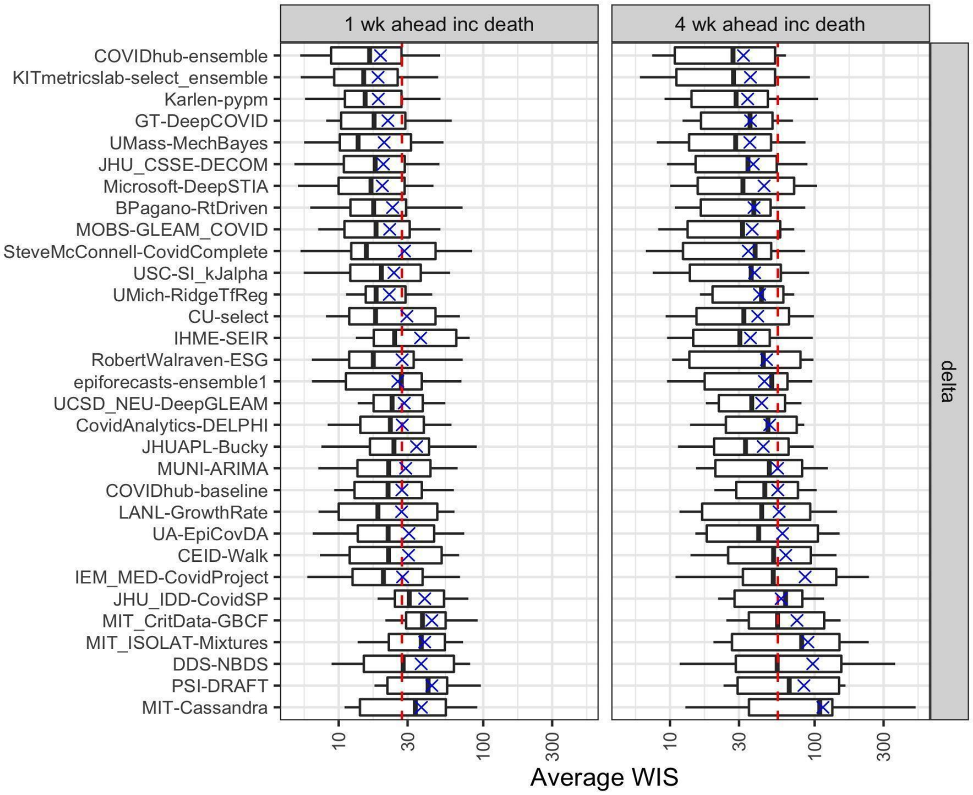


**Fig. S5.** Boxplots of average WIS (shown on log scale), by model and forecast horizon and phase. The boxplots represent the median and interquartile range of the model's weekly average WIS aggregated across locations. The baseline median is shown with a dashed red line, and a team's average WIS is shown with a blue "X". The one week ahead forecasts (left) are more accurate for every model than the 4 week ahead forecasts (right). Models are ordered along the x-axis by their relative WIS within each phase. Based on this aggregation, the ensemble had the lowest relative WIS during only the winter phase.

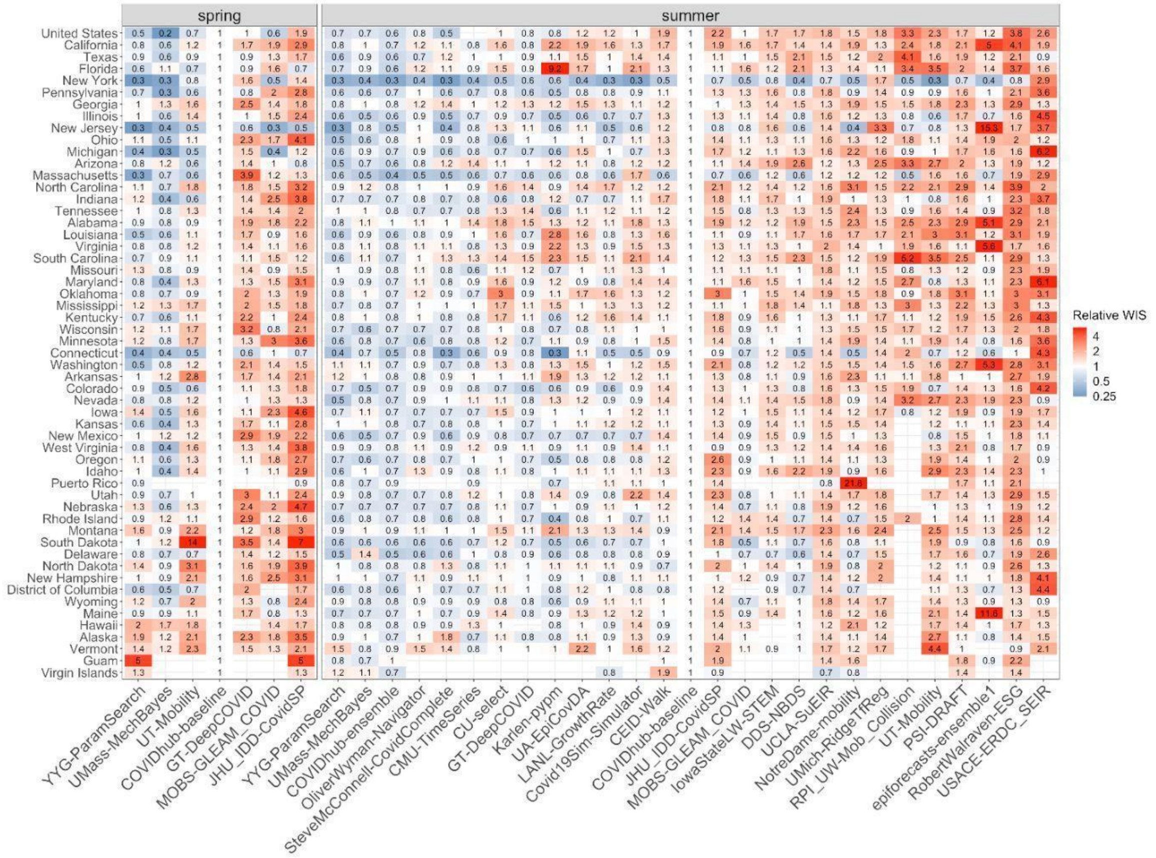
spring
summer
Relative WIS
4
2
1
0.5
0.25
United States
California
Texas
Florida
New York
Pennsylvania
Georgia
Illinois
New Jersey
Ohio
Michigan
Arizona
Massachusetts
North Carolina
Indiana
Tennessee
Alabama
Louisiana
Virginia
South Carolina
Missouri
Maryland
Oklahoma
Mississippi
Kentucky
Wisconsin
Minnesota
Connecticut
Washington
Arkansas
Colorado
Nevada
Iowa
Kansas
New Mexico
West Virginia
Oregon
Idaho
Puerto Rico
Utah
Nebraska
Rhode Island
Montana
South Dakota
Delaware
North Dakota
New Hampshire
District of Columbia
Wyoming
Maine
Hawaii
Alaska
Vermont
Guam
Virgin Islands
YYG-ParamSearch
UMass-MechBayes
UT-Mobility
COVIDhub-baseline
GT-DeepCOVID
MOBS-GLEAM_COVID
JHU_IDD-CovidSP
YYG-ParamSearch
UMass-MechBayes
COVIDhub-ensemble
OliverWyman-Navigator
SteveMcConnell-CovidComplete
CMU-TimeSeries
CU-select
GT-DeepCOVID
Karlen-pypm
UA-EpiCovDA
LANL-GrowthRate
Covid19Sim-Simulator
CEID-Walk
COVIDhub-baseline
JHU_IDD-CovidSP
MOBS-GLEAM_COVID
IowaStateLW-STEM
DDS-NBDS
UCLA-SuEIR
NotreDame-mobility
UMich-RidgeTfReg
RPI_UW-Mob_Collision
UT-Mobility
PSI-DRAFT
epiforecasts-ensemble1
RobertWalraven-ESG
USACE-ERDC_SEIR

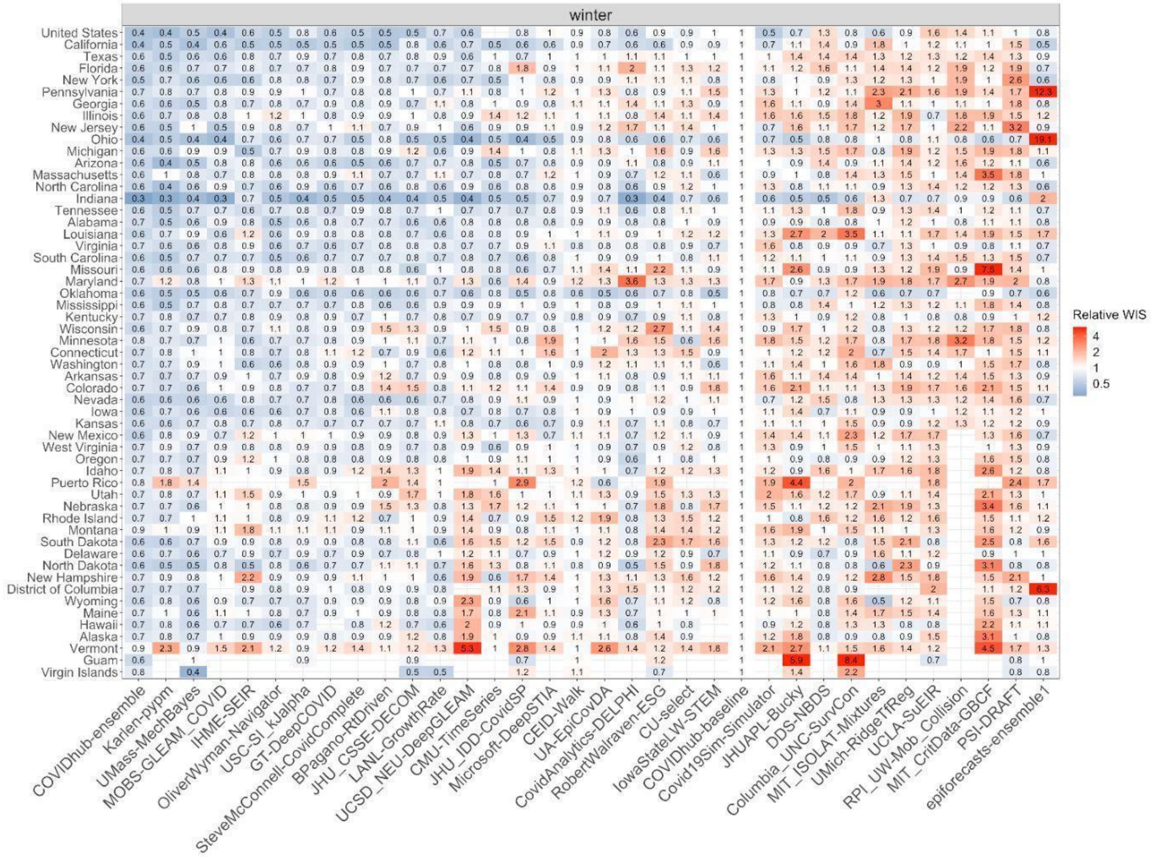
winter
Relative WIS
4
2
1
0.5
COVIDhub-ensemble
Karlen-pypm
UMass-MechBayes
MOBS-GLEAM_COVID
IHME-SEIR
OliverWyman-Navigator
USC-SI_kJalpha
GT-DeepCOVID
SteveMcConnell-CovidComplete
BPagano-RtDriven
JHU_CSSE-DECOM
LANL-GrowthRate
UCSD_NEU-DeepGLEAM
CMU-TimeSeries
JHU_IDD-CovidSP
Microsoft-DeepSTIA
CEID-Walk
UA-EpiCovDA
CovidAnalytics-DELPHI
RobertWalraven-ESG
CU-select
IowaStateLW-STEM
COVIDhub-baseline
Covid19Sim-Simulator
JHUAPL-Bucky
DDS-NBDS
Columbia_UNC-SurvCon
MIT_ISOLAT-Mixtures
UMich-RidgeTfReg
UCLA-SuEIR
RPI_UW-Mob_Collision
MIT_CritData-GBCF
PSI-DRAFT
epiforecasts-ensemble1

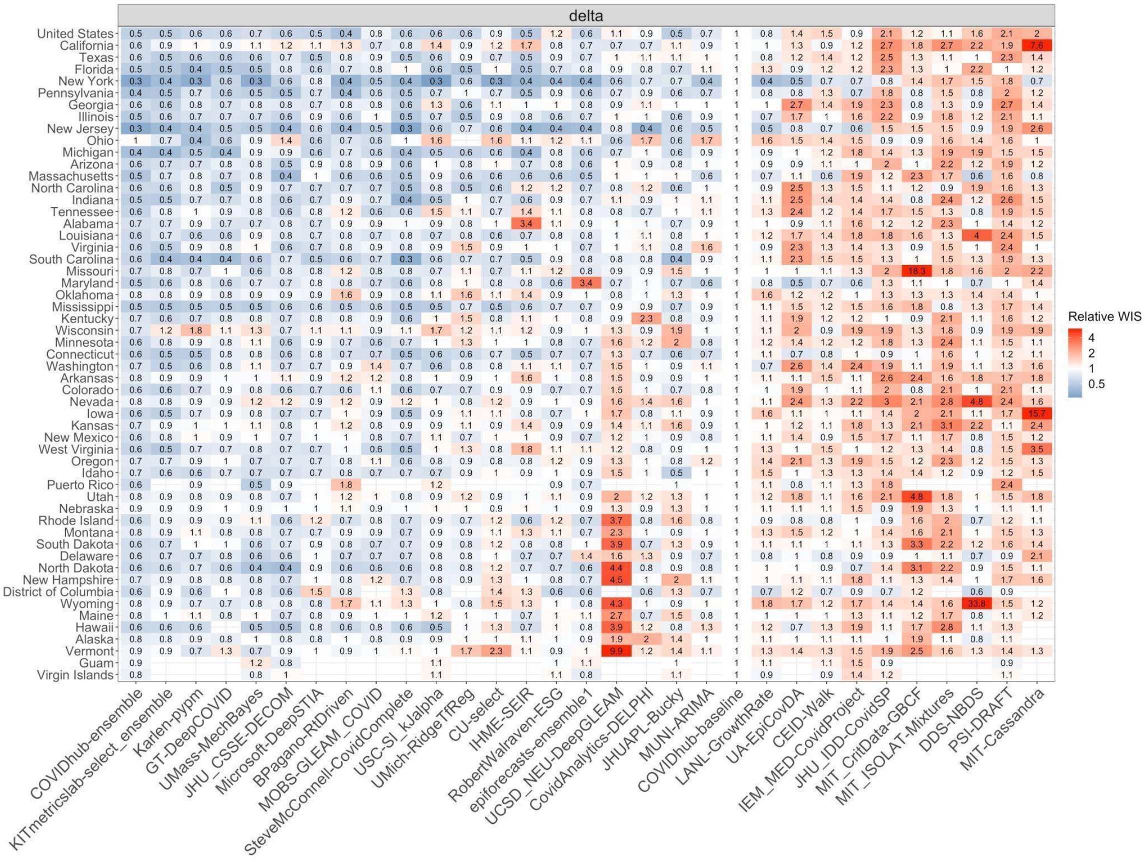


**Fig. S6.** Relative WIS by location for each model across all horizons stratified by pandemic phase. The value in each box represents the relative WIS calculated from 1- to 4-week ahead targets available for a model at each location. Points are colored based on the relative WIS compared to the baseline model ($\theta^{*}_{m}$, see Methods). Blue boxes represent teams that outperformed the baseline and red boxes represent teams that performed worse than the baseline, with darker hues representing performance further away from the baseline. Teams on the x-axis are listed in order from the highest to lowest relative WIS values within each phase. YYG-ParamSearch achieved the lowest average WIS in the spring and summer and the COVIDhub-ensemble achieved the lowest average WIS in the winter. The COVIDhub-ensemble performed at least as well as the baseline in every location.

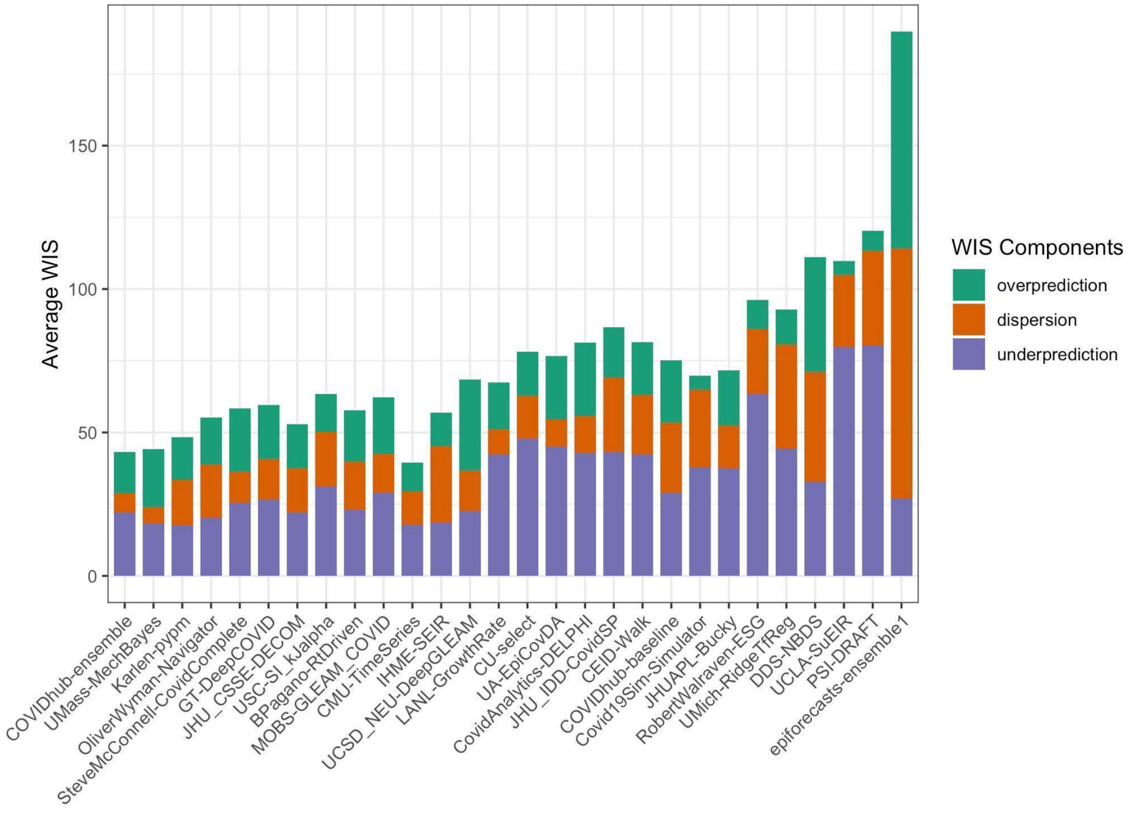


**Fig. S7.** Decomposition of average WIS scores into underprediction, overprediction, and dispersion, aggregated over locations, horizons, and submission weeks. The sum value of these average metrics add up to the average weighted interval score. Models on the x-axis are ordered by relative WIS values (Table 1). The WIS and relative WIS do not follow the same ordering because the WIS values shown below are not adjusted for prediction difficulty across submission weeks and locations.

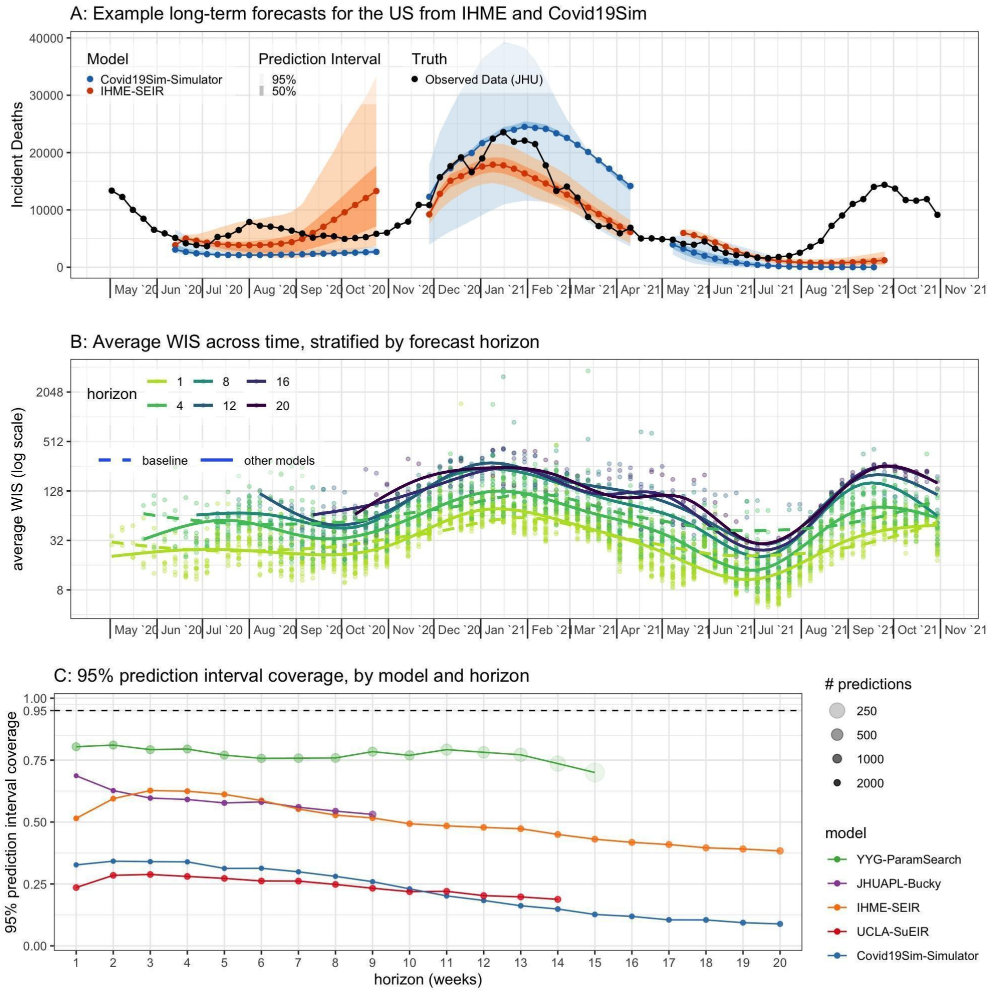


**Fig. S8.** Evaluation of long-range forecast performance. (A) Example 20-week-ahead probabilistic forecasts submitted in early June, and late November 2020, and early May 2021. (B) Points show values of average WIS for specific models and target forecast week across all states. The solid line shows the smooth trend in average WIS across all non-baseline models, and the dashed line shows the trend for the baseline model (horizons 1 and 4 only). Lines are colored by horizon, with darker lines indicating forecasts targeting weeks further in the future. Across all weeks, average WIS tends to be about twice as high for 4-week ahead as it is for 1-week ahead forecasts. For later weeks, when forecasts at all horizons are able to be evaluated, forecasts for horizons above 8 weeks tend to have about double the average WIS as was achieved at a 4-week ahead horizon for these models. (C) 95% prediction interval coverage rates across horizons for a subset of sixfive models that consistently submitted for more than 8 weekly horizons. Coverage rates for 8- through 20-week ahead horizons were all below the nominal 95%. The horizontal dashed line shown at 0.95 indicates the expected coverage rate. The size of points indicates the number of predictions the coverage rates are based on: smaller points indicate more observations and therefore less variance in the estimated coverage rate.

**Fig S9.**

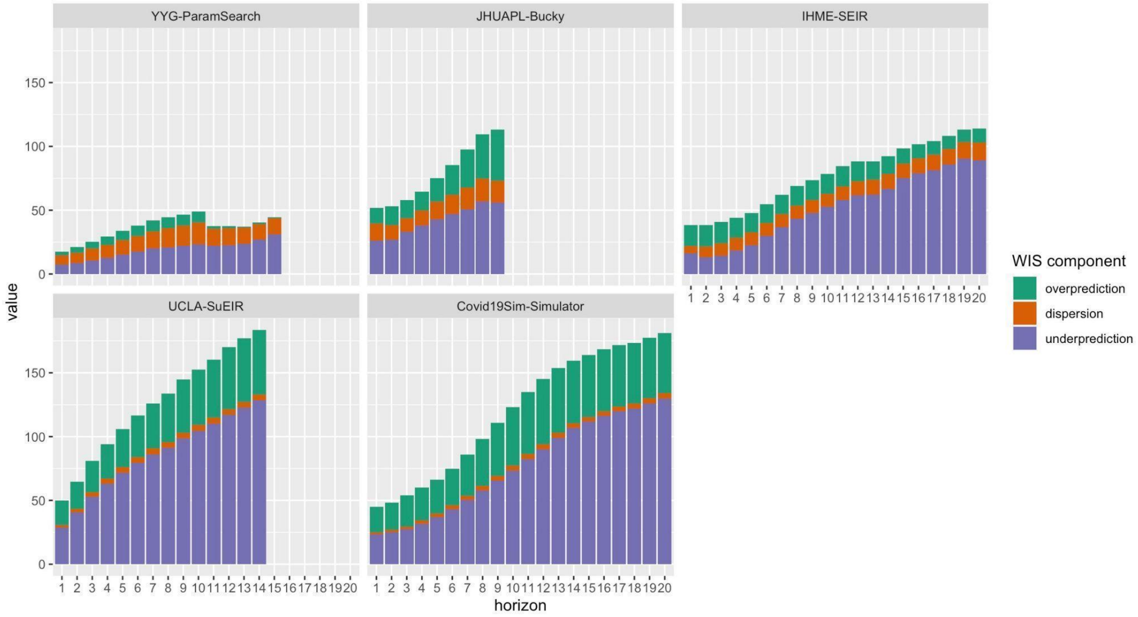


**Fig S9.** Average WIS components by horizon. For models that had over 100 forecasts for horizons greater than 8 weeks, the average WIS components of dispersion, underprediction and overprediction are shown. Average WIS in general increases with horizon. Underprediction tends to increase proportionally more than other components.

**Table S1.** List of models evaluated, including sources for case, hospitalization, death, demographic and mobility data when used as inputs for the given model. We evaluated 28 models contributed by 26 teams. The COVIDhub team submitted two models including the baseline model and the ensemble model. A brief description is included for each model, with a reference where available. The last column indicates whether the model made assumptions about how and whether social distancing measures were assumed to change during the period for which forecasts were made.

| | Data Sources Included | | | | | Model Information | |
|---|---|---|---|---|---|---|---|
| **Team-Model** | **Cases** | **Hosp.** | **Deaths** | **Demog.** | **Mob.** | **Description** | **Assumes social distancing measures change in the future** |
| BPagano-RtDriven | J | | J | | | Death-based SIR model that uses the change history of the Covid-19 effective transmission rate to forecast deaths and cases. | No |
| CEID-Walk | | | J | | | Random walk model starting from the most recent observation with a dispersion based on the spread of the last 5 observations (3) | No |
| CMU-TimeSeries | J | | J | | | A basic autoregressive-type time series model fit using case counts and deaths as features | No |
| Covid19Sim-Simulator | J | CTP | J | | | SEIR model accounting for undiagnosed infections | No |
| COVIDAnalytics-DELPHI | J | | J | | | SEIR model augmented with underdetection and interventions. | Yes |
| COVIDhub-baseline | | | J | | | Median prediction at all future horizons is equal to the most recent observed incidence | No |
| COVIDhub-ensemble | | | | | | Unweighted average or median of submitted forecasts to the COVID-19 Forecast Hub (4) | No |

| | | | | | | | |
|---|---|---|---|---|---|---|---|
| CU-select | J, UF | CTP, HHS | J, UF | Cen | SG, Cen | Metapopulation county-level SEIR model (5-7) | Yes |
| DDS-NBDS | J | | J | | | Negative binomial distribution based generalized linear dynamical system | No |
| epiforecasts-ensemble1 | J | | J | | | Mean ensemble of three models: an Rt-based forecast, a timeseries forecast using deaths only and a timeseries forecast using deaths and cases | No |
| GT-DeepCOVID | CTP | CTP, HHS, CN | J | | G,A | Data-driven approach based on deep learning for forecasting mortality and hospitalizations (8) | No |
| IHME-SEIR[a] | J, CTP | CTP, HHS | J, CTP | GBD | SG, G, USDT, FB | Ensemble spline model to estimate past infections combined with covariate-driven deterministic SEIR model | Yes |
| JHU_CSSE-DECOM | J | | J | Cen | SG | County-level, empirical machine learning model driven by epidemiological, mobility, demographic, and behavioral data. | No |
| JHU_IDD-CovidSP[c] | J,UF | | J,UF | Cen | Cen | Metapopulation model with commuting, nonpharmaceutical interventions, and stochastic SEIR disease dynamics (9) | No |
| JHUAPL-Bucky | J | HHS | J | Cen | SG, PIQ | Spatial compartment model using public mobility data and local parameters | Yes |
| Karlen-pypm | J | HHS | J | | | Finite time difference equations implemented as a general-purpose population modelling framework (10, 11) | No |
| LANL-GrowthRate[d] | J | | J | | | Statistical dynamical growth model accounting for population susceptibility (12) | No |

| | | | | | | | |
|---|---|---|---|---|---|---|---|
| MOBS-GLEAM_COVID | J | HHS | J | Cen | G | Metapopulation, age-structured SLIR model with mobility and nonpharmaceutical interventions (13) | Yes |
| OliverWyman-Navigator | J | | J | Cen | | Compartmental formulation with non-stationary transition rates | Blended. (No for immediate term up to next 3 weeks. Yes for longer term.) |
| PSI-DRAFT | | | J | Cen | | Age-stratified compartmental SEIRX model with time-dependent reproduction number | No |
| RobertWalraven-ESG | J | | J | | | Multiple skewed gaussian mathematical fit | No |
| SteveMcConnell-CovidComplete | CTP | | J, CTP | Cen | | Multiple proxy-based forecast models with positive tests and past deaths used as proxies for future deaths; ongoing accuracy evaluation of each model; voting algorithms based on past performance used to select specific forecast models each week, selected state by state; ; most forecasts are error-corrected based on errors in past forecasts | No |
| UA-EpiCovDA[e] | CTP, J | | CTP, J | | | SIR mechanistic model with data assimilation (14, 15) | No |
| UCLA-SuEIR | J | CTP | J | | | SEIR model variant considering both untested and unreported cases | Yes |
| UCSD_NEU-DeepGLEAM | J | HHS | J | Cen | G | Combines the signal of a discrete stochastic epidemic computational model with a deep learning spatiotemporal forecasting framework (16, | Yes |

| | | | | | | | |
|---|---|---|---|---|---|---|---|
| | | | | | | 17) | |
| UMass-MechBayes | J | | J | | | Bayesian compartmental model with observations on incident case counts and incident deaths (18) | No |
| UMich-RidgeTfReg[f] | J | | J | | G | Ridge regression model using confirmed case and death reports to generate predictions | No |
| USC-SIkJalpha | J | HHS | J | | | Models temporally varying infection, death, and hospitalization rates. Learning is performed by reducing the problem to multiple simple linear regression problems. True susceptible population is identified based on reported cases, whenever mathematically possible (19, 20) | No |

A = Apple mobility (https://covid19.apple.com/mobility), Cen = US Cen (https://www.census.gov/), CN = Coronavirus Disease 2019 (COVID-19)-Associated Hospitalization Surveillance Network (COVID-NET) (https://www.cdc.gov/coronavirus/2019-ncov/covid-data/covid-net/purpose-methods.html), CTP = COVID Tracking Project (https://covidtracking.com/), DL= Descartes Labs (https://github.com/descarteslabs/DL-COVID-19), FB = Facebook (https://visualization.covid19mobility.org/), G = Google mobility (https://www.google.com/covid19/mobility/), GBD = Global Burden of Disease project (http://www.healthdata.org/gbd/2019), HHS = Health and human services hospitalizations (https://protect-public.hhs.gov/pages/covid19-module), J = JHU CSSE (https://github.com/CSSEGISandData/COVID-19) (21), NYT = New York Times (https://github.com/nytimes/covid-19-data), SEIR = Susceptible-Exposed-Infectious-Recovered compartmental model, SG = SafeGraph mobility (https://www.safegraph.com/), SIR = Susceptible-Infectious-Recovered compartmental model, UF = USA Facts (https://usafacts.org/visualizations/coronavirus-covid-19-spread-map/), USDT = U.S. Department of Transportation Bureau of Transportation Statistics (https://www.transportation.gov/connect/available-datasets)

[a]The IHME-SEIR model on 2020-06-24 switched from curve fitting for past infections and SEIR model for infection projections to using an ensemble spline model to estimate past infections combined with covariate-driven deterministic SEIR model
[b]The IowaStateLW-STEM model on 2020-07-27 switched from using the NYT data to JHU CSSE data and started incorporating mobility data.
[c]The JHU_IDD-CovidSP model on 2020-12-14 switched to using JHU CSSE data only for cases and deaths.
[d]The LANL-GrowthRate model on 2020-10-28 switched from a Bayesian hierarchical approach to share information between states to fitting each state separately for improved computational time.
[e]The UA-EpiCovDA model on 2020-07-05 switched the way the initial conditions were being estimated. After March 8, 2021, forecasts were updated using JHU CSSE instead of CTP.
[f]The UMich-RidgeTfReg model on 2020-11-30 started to incorporate social mobility data.

**Supplemental Table 2:** Summary of models that contributed to the ensemble forecast but were not individually evaluated due to not having enough eligible submissions during the evaluation period.

| | Data Sources Included | | | | | Model Information | |
|---|---|---|---|---|---|---|---|
| **Team-Model** | **Cases** | **Hosp.** | **Deaths** | **Demog.** | **Mob.** | **Description** | **Assumes social distancing measures change in the future (data source)** |
| Alpert-pwllnod | | | J | | | Piecewise Log Linear model using policy change dates. | No |
| CovidActNow-SEIR_CAN | NYT | | NYT | | | SEIR model | No |
| Columbia_UNC-SurvCon | J | | J | | | Survival-convolution model with piecewise transmission rates that incorporates latent incubation period and provides time-varying effective reproductive number. | No |
| Google_Harvard-CPF | J | CTP | J | BQ | DL | Extended SEIR model with hospitalization compartments and trainable encoders that process static and time-varying covariates to extract information from. Trained in an end-to-end way with partial teacher forcing. | Yes (CHC) |
| GT_CHHS-COVID19 | GA DPH, NC DHHS | | GA DPH, NC DHHS | Cen | Cen, SG, SL | Agent-based simulation disease spread model assuming heterogeneous population mixing to predict the spread pattern geographically over time (22, 23) | Yes |
| IEM_MED-CovidProject | J | | J | | | SEIR model projections using MCMC to find best parameters to fit actual data. | No |
| IowaStateLW-STEM[b] | J,NYT | | J,NYT | Cen | US DT | Nonparametric space-time disease transmission model (24) | No |
| JCB-PRM | J | | J | | | Deterministic model built on observations of macro-level societal and political responses to COVID measured only in terms of infections and deaths. | Yes |
| LNQ-ens1 | | | | J | | County-level ensemble of boosted tree and neural net models | No |
| MIT-Cassandra | J | | J | Cen | G | Ensemble model combining four types of models (minimum representation | No |

| | | | | | | | |
|---|---|---|---|---|---|---|---|
| | | | | | | learning, nearest neighbors matching on time-series, deep learning and epidemiology) to forecast deaths (25) | |
| MIT_CritData-GBCF | J | | J | Cen | PIQ | Gradient boosted regressor with hyperparameter optimization that uses prior COVID-19 cases and deaths as well as static and time-varying county-level covariates. Forecasts at county-level and aggregates to state and national level. | No |
| MITCovAlliance-SIR | NYT | | NYT | Cen, CDC, CL, UM | SG | SIR model trained on public health regions. SIR parameters are functions of static demographic and time-varying mobility features. A two-stage approach that first learns the magnitude of peak infections (26) | No |
| MIT_ISOLAT-Mixtures | J | | | J | | Non-mechanistic, non-parametric model based on representing time series as a sum of bell curves. | No |
| Microsoft-DeepSTIA | J | CTP | J | | G | A hierarchical spatial-temporal forecasting model that not only follows the time-series trends but also takes into consideration the spatial correlations among different administrative regions. | Yes |
| MSRA-DeepST | J | CTP | J | | G | Deep spatio-temporal network with knowledge-based SEIR as a regularizer under the assumption of spatio-temporal process in pandemic of different regions. | Yes |
| MUNI-ARIMA | | | J | | | ARIMA model with outlier detection fitted to transformed weekly aggregated series | No |
| NotreDame-FRED | NYT | | NYT | | | Agent-based model developed for influenza with parameters modified to represent the natural history of COVID-19. | Yes (IHME COVID-19 health service utilization forecasting Team) |
| NotreDame-mobility | CTP | | J | | G,A | Ensemble of nine models that are identical except that they are driven by different mobility indices from Apple and Google. Underlying deterministic, SEIR-like model. | No |
| USACE-ERDC_SEIR | J,UF | CTP | J,UF | | | SEIR model with additional compartments for unreported infections and isolated individuals (27) | No |
| QJHong-Encounter | J | CTP | J | | | SEIR model using encounter density to predict reproductive number | No |

| | | | | | | | |
|---|---|---|---|---|---|---|---|
| RPI_UW-Mob_Collision | | | J | | G | A mobility-informed simplified SIR model motivated by collision theory. | No |
| SigSci-TS | J | | J | | | Time series forecasting using ARIMA for case forecasts and lagged cases for death forecasts. | No |
| SWC-TerminusCM | CTP | CTP | CTP | | | Mechanistic compartmental model using disease parameter estimates from literature and Bayesian inference. | Yes |
| UCM_MESALab-FoGSEIR | J | | J | | G | Modification of integer order SEIR model considering fractional integrals. Considers the age structure and reopening intervention to minimize infections and deaths. | Yes |
| UCSB-ACTS | J | CTP | J | | | Data-driven machine learning model that makes predictions by referring to other regions with similar growth patterns and assuming similar development will take place in the current region. | No |
| UpstateSU-GRU | J | | Cen, KFF, BRFSS | J | G | Recurrent neural network seq2seq model with the Gated recurrent units (28) | Yes |
| UT-Mobility | | | J | | SG | Bayesian multilevel negative binomial regression model | No |
| Wadhwani_AI-BayesOpt | J | | J | | | Model-agnostic Bayesian optimization ("BayesOpt") approach for learning the parameters of an SEIR-like compartmental model from observed data. | No |
| YYG-ParamSearch | | | J | Cen | | SEIR model with a machine learning layer (29) | Yes |

BQ = Bigquery public datasets (https://cloud.google.com/bigquery/public-data),BRFSS = Behavioral Risk Factor Surveillance System (https://www.cdc.gov/brfss/data_documentation/index.htm), Cen = US Cen (https://www.census.gov/), CHC = COVID Healthcare Coalition https://c19hcc.org/resources/npi-dashboard/),
CL = Claritas (https://www.claritascreative.com/covid19), CN = Coronavirus Disease 2019 (COVID-19)-Associated Hospitalization Surveillance Network (COVID-NET) (https://www.cdc.gov/coronavirus/2019-ncov/covid-data/covid-net/purpose-methods.html), CTP = COVID Tracking Project (https://covidtracking.com/), DL= Descartes Labs (https://github.com/descarteslabs/DL-COVID-19), G = Google mobility (https://www.google.com/covid19/mobility/), GA DPH = Georgia Department of Public Health (https://dph.georgia.gov/covid-19-daily-status-report), HHS = Health and human services hospitalizations (https://protect-public.hhs.gov/pages/covid19-module), J = JHU CSSE (https://github.com/CSSEGISandData/COVID-19)(24) , KFF = regional health index from 2019 Kaiser Family Foundation Survey (https://www.kff.org/report-section/ehbs-2019-summary-of-findings/), MMODS = Multi-modal outbreak decision support scenarios (https://midasnetwork.us/mmods/), NC DHHS = NC Department of Health and Human Services (https://covid19.ncdhhs.gov/dashboard), NYT = New York Times (https://github.com/nytimes/covid-19-data), PIQ = Place IQ

(https://github.com/COVIDExposureIndices/COVIDExposureIndices), Rt = time-varying reproductive number, SEIR = Susceptible-Exposed-Infectious-Recovered compartmental model, SG = SafeGraph mobility (https://www.safegraph.com/), SIR = Susceptible-Infectious-Recovered compartmental model, SL = StreetLight (https://www.streetlightdata.com/), UM = University of Michigan Health and Retirement Study (https://hrs.isr.umich.edu/data-products)

**Supplemental Table 3:** Sensitivity analysis of relative WIS calculations. We computed the relative WIS (rel WIS, $\theta^{*}_{m}$) across three different time periods and using two different inclusion criteria, to assess the robustness of the inclusion criteria applied for model selection. The results show that the values of relative WIS and the ordering of models according to this metric were not strongly sensitive to whether models with smaller numbers of available forecasts were included in the computation of relative WIS. (The "% max obs" column shows the percentage of the maximum possible scores that a given model made.) Some models showed differences in relative WIS when different weeks were included, which is to be expected if models performed better during different phases of the pandemic.

| | | Exclusion criteria applied | Sensitivity 1 | Sensitivity 2 |
|---|---|---|---|---|
| time period evaluated | | EW18-2020 - EW17-2021 | EW18-2020 - EW17-2021 | EW18-2020 - EW17-2021 |
| Inclusion criteria | | >= 32 weeks submitted (60%) | >= 37 weeks submitted (70%) | >= 47 weeks (89%) |
| model | % Max Obs | rel WIS | rel WIS | rel WIS |
| CEID-Walk | 69.38 | 0.95 | - | - |
| CMU-TimeSeries | 76.23 | 0.79 | 0.79 | - |
| Covid19Sim-Simulator | 86.41 | 1.01 | 1.00 | - |
| COVIDhub-baseline | 100.00 | 1.00 | 1.00 | 1.00 |
| COVIDhub-ensemble | 88.33 | 0.61 | 0.61 | 0.62 |
| CU-select | 84.44 | 0.96 | 0.96 | - |
| DDS-NBDS | 72.78 | 1.16 | 1.17 | - |
| epiforecasts-ensemble1 | 68.34 | 3.88 | 3.81 | - |
| GT-DeepCOVID | 84.44 | 0.77 | 0.78 | 0.77 |
| IHME-SEIR | 67.45 | 0.77 | 0.77 | - |
| IowaStateLW-STEM | 75.47 | 1.03 | 1.03 | - |
| JHU_IDD-CovidSP | 93.57 | 0.88 | 0.89 | 0.92 |
| JHUAPL-Bucky | 63.09 | 1.10 | - | - |
| Karlen-pypm | 76.66 | 0.66 | 0.67 | - |
| LANL-GrowthRate | 86.39 | 0.80 | 0.80 | - |
| MOBS-GLEAM_COVID | 99.92 | 0.81 | 0.82 | 0.83 |
| OliverWyman-Navigator | 88.10 | 0.70 | 0.70 | 0.71 |

| | | | | |
|---|---|---|---|---|
| PSI-DRAFT | 80.44 | 1.47 | 1.49 | - |
| RobertWalraven-ESG | 80.26 | 1.26 | 1.28 | - |
| RPI_UW-Mob_Collision | 40.42 | 1.34 | 1.36 | - |
| SteveMcConnell-CovidComplete | 66.97 | 0.74 | - | - |
| UA-EpiCovDA | 84.44 | 0.93 | 0.93 | - |
| UCLA-SuEIR | 79.65 | 1.28 | 1.28 | - |
| UCSD_NEU-DeepGLEAM | 63.09 | 0.81 | - | - |
| UMass-MechBayes | 96.11 | 0.61 | 0.62 | 0.62 |
| UMich-RidgeTfReg | 66.35 | 1.30 | 1.31 | - |
| UT-Mobility | 69.46 | 2.66 | 2.57 | - |

**Supplemental Table 4**: Sensitivity analysis examining the impact of excluding data anomalies (outlying observations, or forecasts made from revised data) on the calculations of relative WIS, relative MAE and prediction interval coverage for the evaluation period from EW17-2020 through EW16-2021. In general, the metrics do not show large differences based on including or not these anomalous observations in the evaluation.

| | Full analysis for EW16-2020 - EW17-2021 | | | | | Sensitivity analysis (no anomalies) for EW16-2020 - EW17-2021 | | | | |
|---|---|---|---|---|---|---|---|---|---|---|
| | | PI Cov | | | | | PI Cov | | | |
| **model** | **N** | **95%** | **50%** | **relWIS** | **relMAE** | **N** | **95%** | **50%** | **relWIS** | **relMAE** |
| CEID-Walk | 7135 | 0.81 | 0.46 | 0.95 | 1.00 | 6608 | 0.82 | 0.47 | 0.95 | 1.01 |
| CMU-TimeSeries | 7840 | 0.72 | 0.39 | 0.79 | 0.80 | 7298 | 0.73 | 0.40 | 0.78 | 0.79 |
| Covid19Sim-Simulator | 8886 | 0.27 | 0.08 | 1.01 | 0.81 | 8326 | 0.28 | 0.08 | 1.03 | 0.82 |
| COVIDhub-baseline | 10284 | 0.84 | 0.44 | 1.00 | 1.00 | 9706 | 0.85 | 0.46 | 1.00 | 1.00 |
| COVIDhub-ensemble | 9084 | 0.87 | 0.47 | 0.61 | 0.66 | 8518 | 0.89 | 0.49 | 0.58 | 0.64 |
| CU-select | 8684 | 0.68 | 0.34 | 0.96 | 0.93 | 8127 | 0.70 | 0.34 | 0.99 | 0.96 |
| DDS-NBDS | 7485 | 0.84 | 0.40 | 1.16 | 1.52 | 6958 | 0.85 | 0.41 | 1.16 | 1.51 |
| epiforecasts-ensemble1 | 7028 | 0.86 | 0.45 | 3.88 | 3.17 | 6505 | 0.87 | 0.45 | 1.55 | 0.93 |
| GT-DeepCOVID | 8684 | 0.82 | 0.37 | 0.77 | 0.85 | 8146 | 0.83 | 0.38 | 0.74 | 0.80 |
| IHME-SEIR | 6937 | 0.64 | 0.27 | 0.77 | 0.80 | 6483 | 0.65 | 0.28 | 0.76 | 0.80 |
| IowaStateLW-STEM | 7761 | 0.44 | 0.18 | 1.03 | 0.92 | 7223 | 0.45 | 0.18 | 1.07 | 0.94 |
| JHU_IDD-CovidSP | 9623 | 0.80 | 0.36 | 0.88 | 0.99 | 9070 | 0.81 | 0.36 | 0.92 | 1.03 |
| JHUAPL-Bucky | 6488 | 0.53 | 0.24 | 1.10 | 1.09 | 5968 | 0.53 | 0.24 | 1.17 | 1.14 |
| Karlen-pypm | 7884 | 0.84 | 0.44 | 0.66 | 0.72 | 7342 | 0.85 | 0.46 | 0.65 | 0.71 |
| LANL-GrowthRate | 8884 | 0.89 | 0.40 | 0.80 | 0.89 | 8322 | 0.900 | 0.40 | 0.79 | 0.89 |
| MOBS-GLEAM_COVID | 10276 | 0.67 | 0.35 | 0.81 | 0.80 | 9698 | 0.68 | 0.36 | 0.81 | 0.79 |
| OliverWyman-Navigator | 9060 | 0.83 | 0.44 | 0.70 | 0.74 | 8498 | 0.84 | 0.46 | 0.68 | 0.72 |
| PSI-DRAFT | 8272 | 0.34 | 0.14 | 1.47 | 1.24 | 7721 | 0.35 | 0.15 | 1.55 | 1.28 |
| RobertWalraven-ESG | 8254 | 0.37 | 0.21 | 1.26 | 1.03 | 7705 | 0.38 | 0.22 | 1.28 | 1.04 |
| RPI_UW-Mob_Collision | 4157 | 0.55 | 0.22 | 1.34 | 1.29 | 3815 | 0.56 | 0.22 | 1.41 | 1.34 |

| | | | | | | | | | | |
|---|---|---|---|---|---|---|---|---|---|---|
| SteveMcConnell-CovidComplete | 6887 | 0.79 | 0.49 | 0.74 | 0.78 | 6353 | 0.80 | 0.50 | 0.72 | 0.76 |
| UA-EpiCovDA | 8684 | 0.64 | 0.33 | 0.93 | 0.88 | 8127 | 0.65 | 0.34 | 0.95 | 0.89 |
| UCLA-SuEIR | 8191 | 0.24 | 0.08 | 1.28 | 1.08 | 7652 | 0.24 | 0.07 | 1.28 | 1.04 |
| UCSD_NEU-DeepGLEAM | 6488 | 0.87 | 0.55 | 0.81 | 0.79 | 5956 | 0.89 | 0.57 | 0.81 | 0.78 |
| UMass-MechBayes | 9884 | 0.94 | 0.56 | 0.61 | 0.66 | 9314 | 0.95 | 0.57 | 0.61 | 0.65 |
| UMich-RidgeTfReg | 6823 | 0.45 | 0.23 | 1.30 | 1.13 | 6518 | 0.47 | 0.24 | 1.36 | 1.17 |
| UT-Mobility | 7143 | 0.67 | 0.30 | 2.66 | 2.45 | 6718 | 0.68 | 0.30 | 2.85 | 2.50 |

**Supplemental Table 5:** Table showing how the current manuscript met the EPIFORGE 2020 checklist of reporting guidelines for epidemiological forecasting studies.

| Section of manuscript | # | Checklist item | Reported on page* |
|---|---|---|---|
| Title / Abstract | 1 | Describe the study as forecast or prediction research in at least the title or abstract | 1, 4 |
| Introduction | 2 | Define the purpose of study and forecasting targets | 5, 6 |
| Methods | 3 | Fully document the methods | 13-18, SI 3-5 |
| Methods | 4 | Identify whether the forecast was performed prospectively, in real-time, and/or retrospectively | 16 |
| Methods | 5 | Explicitly describe the origin of input source data, with references | 14, SI 5 |
| Methods | 6 | Provide source data with publication, or document reasons as to why this was not possible | 14 |
| Methods | 7 | Describe input data processing procedures in detail | 15 |
| Methods | 8 | State and describe the model type, and document model assumptions, including references | 16 |
| Methods | 9 | Make the model code available, or document the reasons why this was not possible | 18 |
| Methods | 10 | Describe the model validation, and justify the approach. | 14-21 |
| Methods | 11 | Describe the forecast accuracy evaluation method used, with justification | 17-18, SI 3-4 |
| Methods | 12 | Where possible, compare model results to a benchmark or other comparator model, with justification of comparator choice | 16, SI 4 |
| Methods | 13 | Describe the forecast horizon, with justification of its length | 14 |
| Results | 14 | Present and explain uncertainty of forecasting results | 8-11 |

| | | | |
|---|---|---|---|
| Results | 15 | Briefly summarize the results in non-technical terms, including a non-technical interpretation of forecast uncertainty | 9-10 |
| Results | 16 | If results are published as a data object, encourage a time-stamped version number | 18 |
| Discussion | 17 | Describe the weaknesses of the forecast, including weaknesses specific to data quality and methods | 12-13 |
| Discussion | 18 | If the research is applicable to a specific epidemic, comment on its potential implications and impact for public health action and decision making | 12 |
| Discussion | 19 | If the research is applicable to a specific epidemic, comment on how generalizable it may be across populations | 13 |

*Page numbers are based on pages in the preprint posted on medRxiv:
**https://www.medrxiv.org/content/10.1101/2021.02.03.21250974v2**
