## Supplementary figures and images for "Evaluation of individual and ensemble probabilistic forecasts of COVID-19 mortality in the US"

### Supplemental File 1

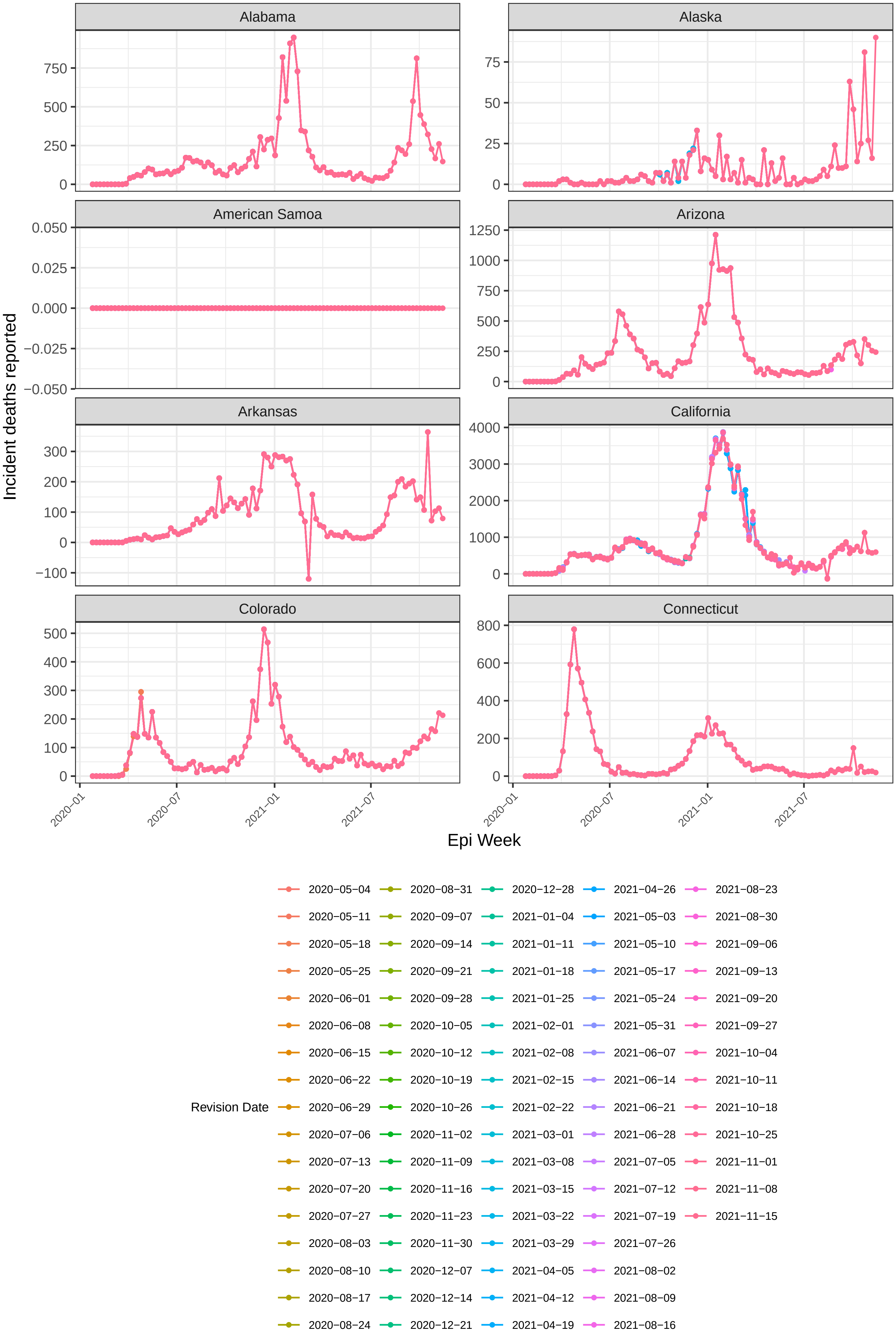

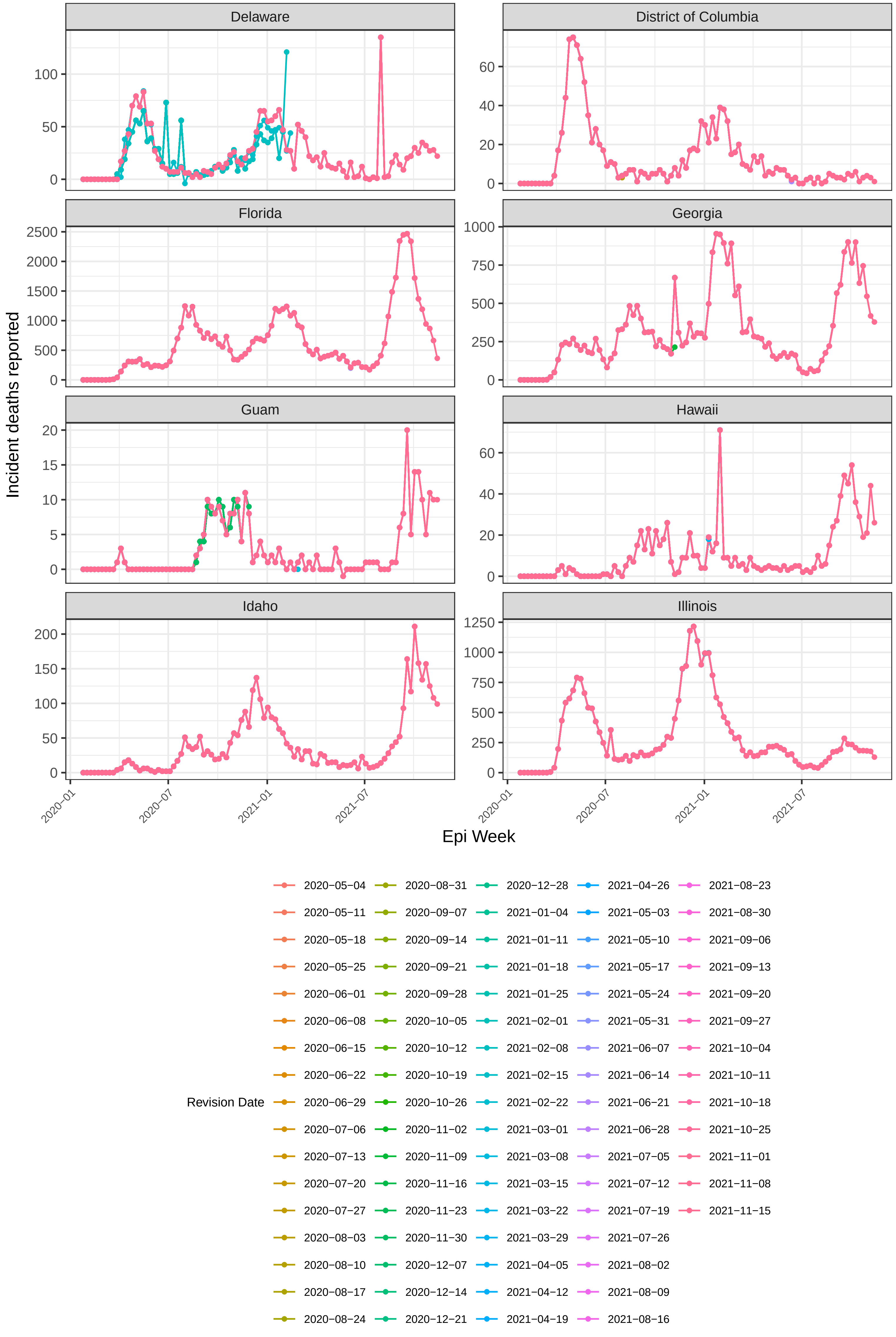

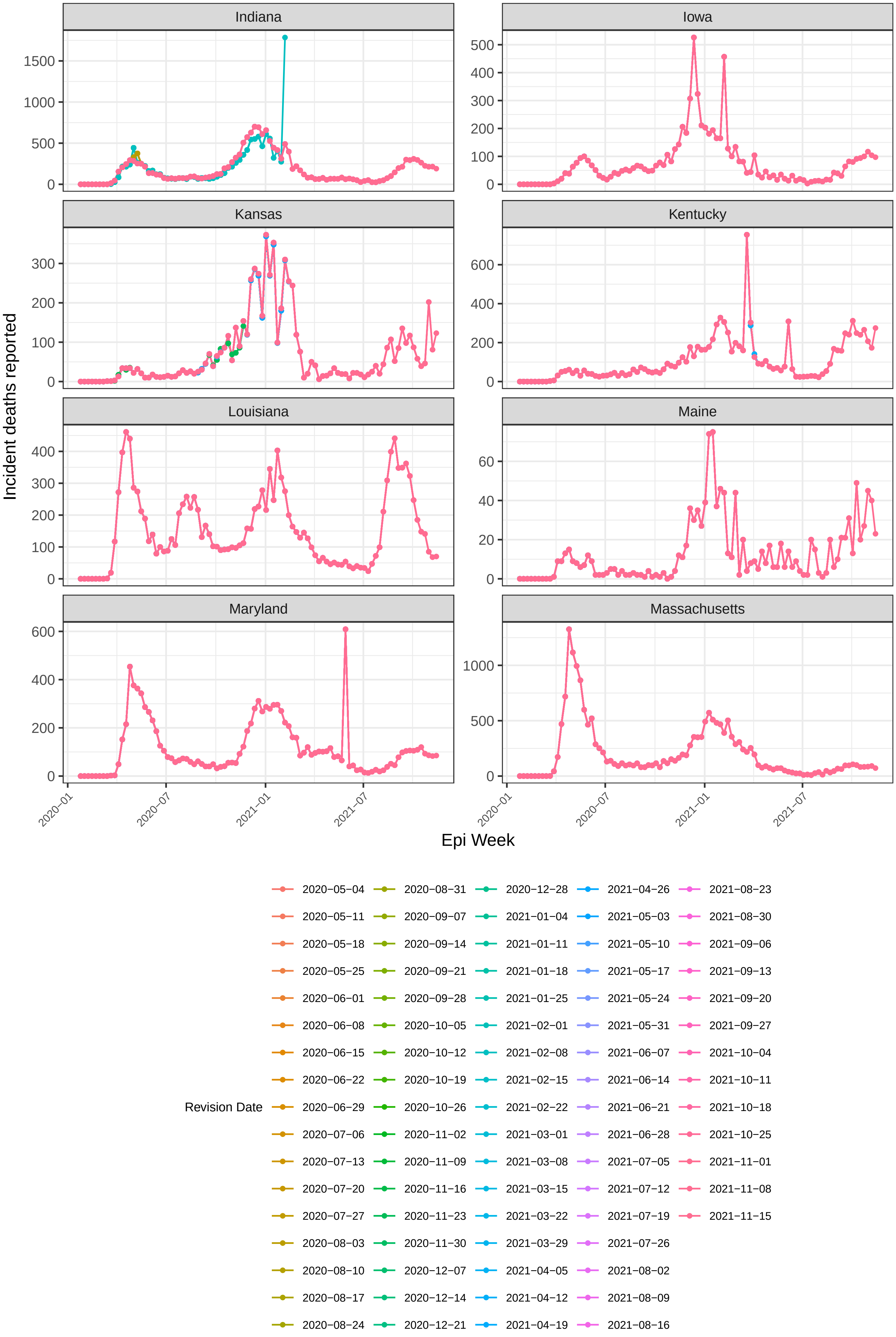

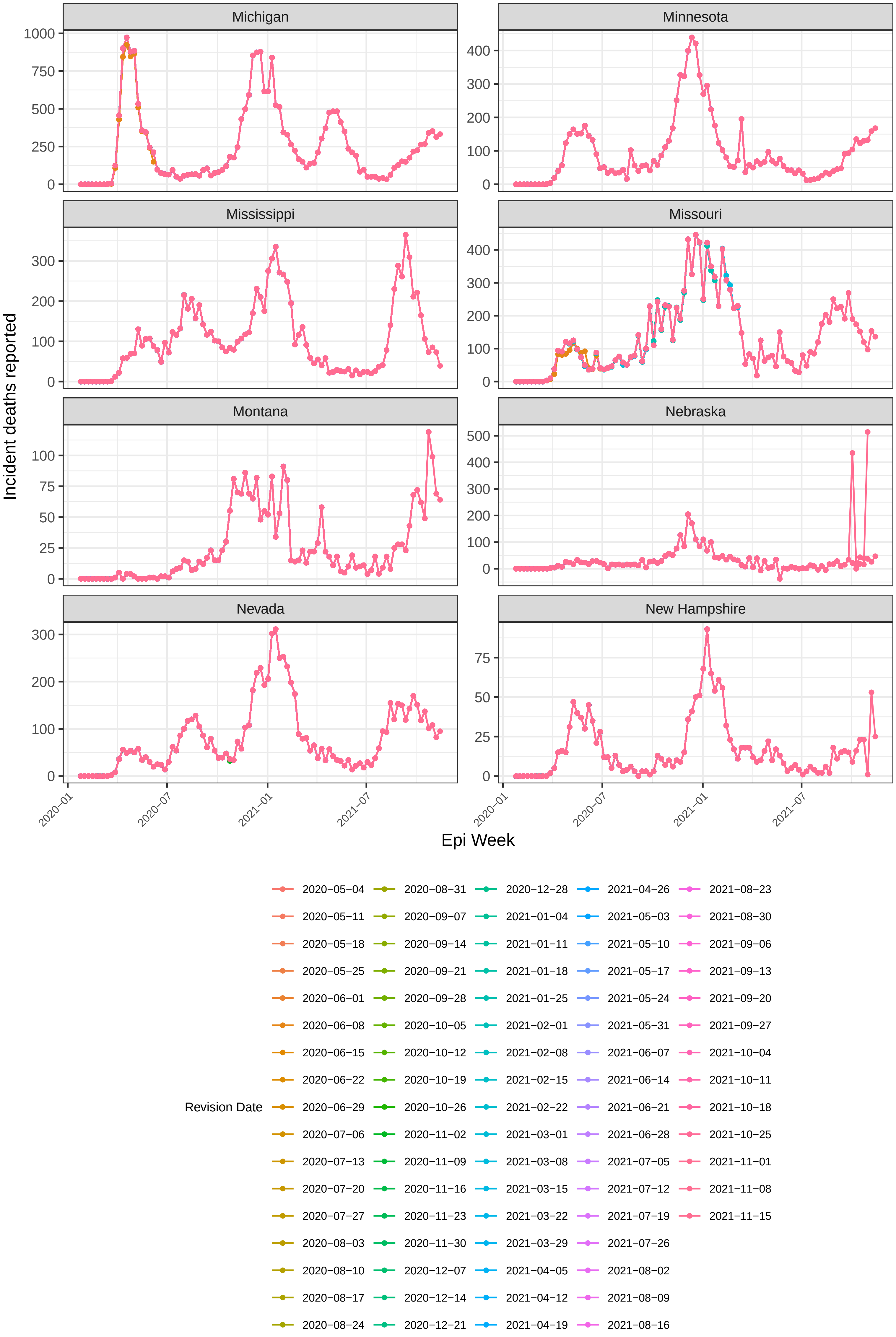

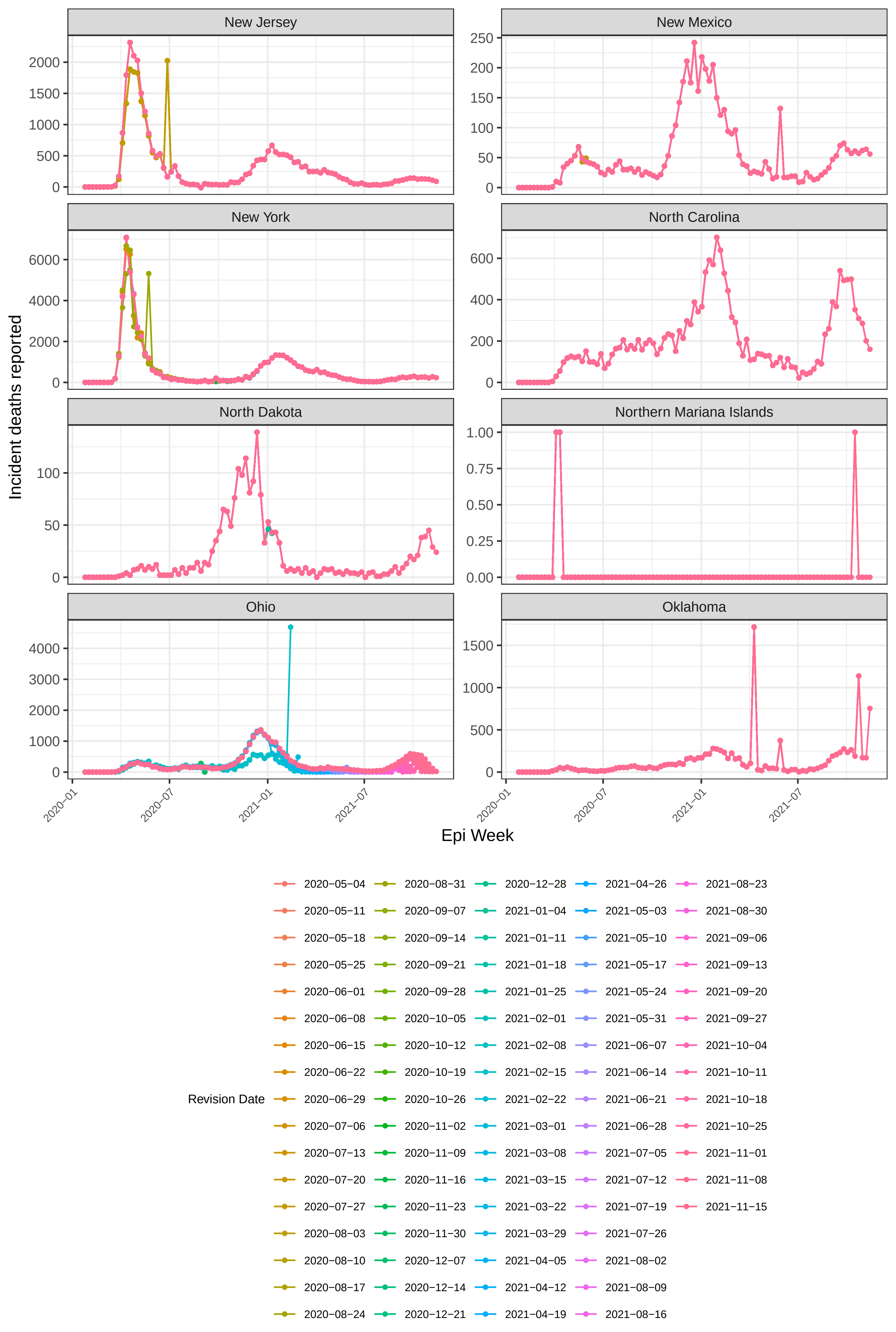

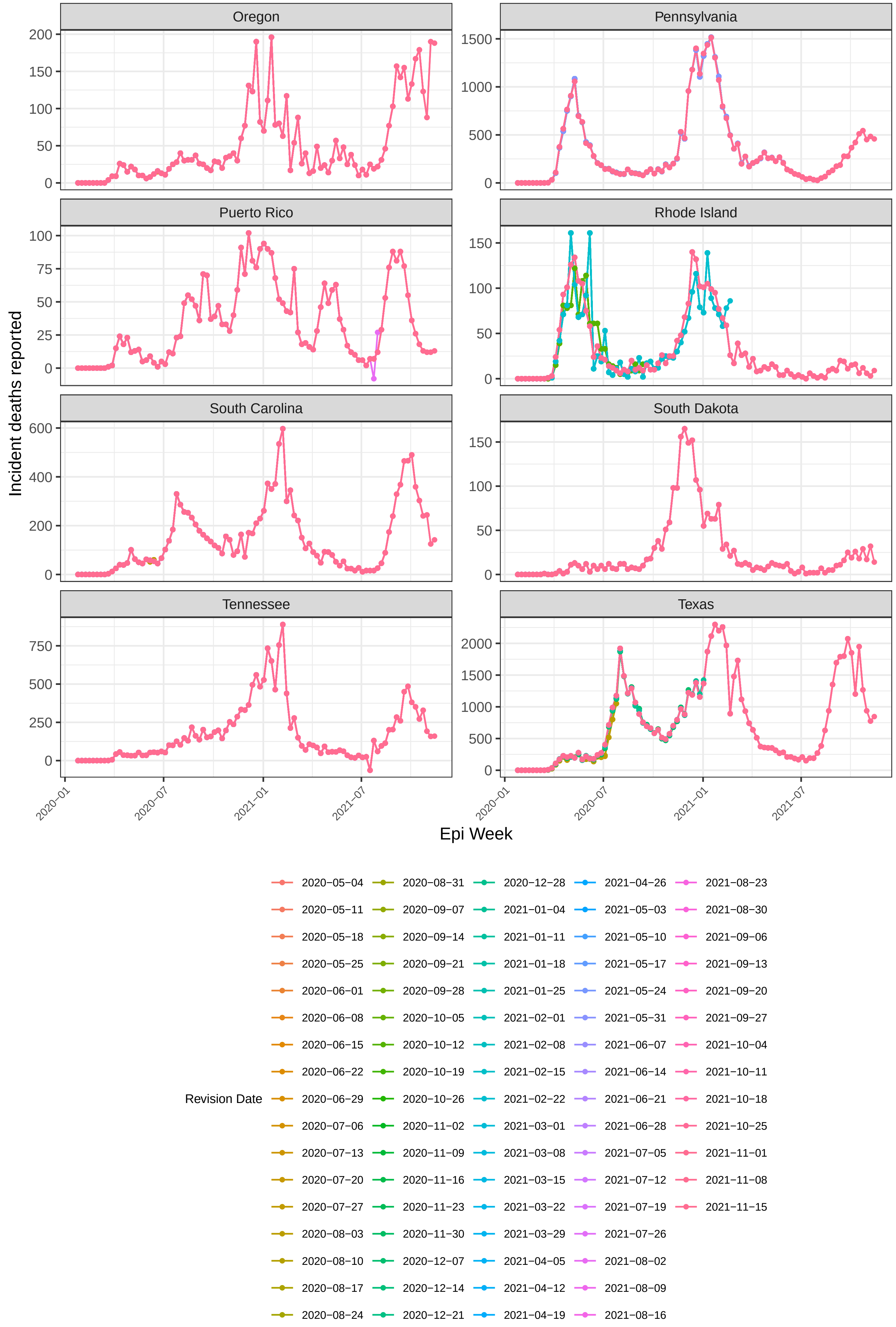

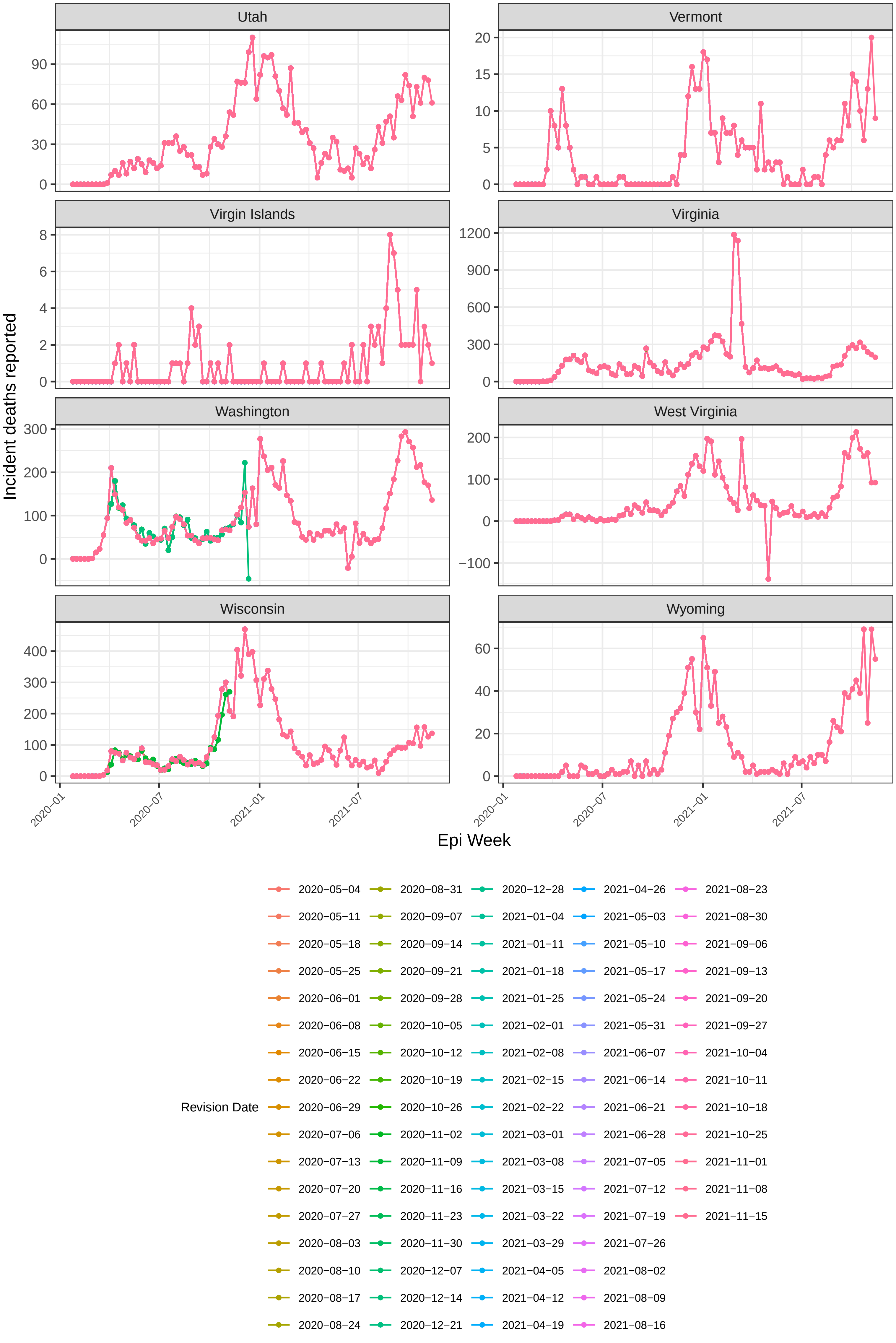

### Supplemental File 2

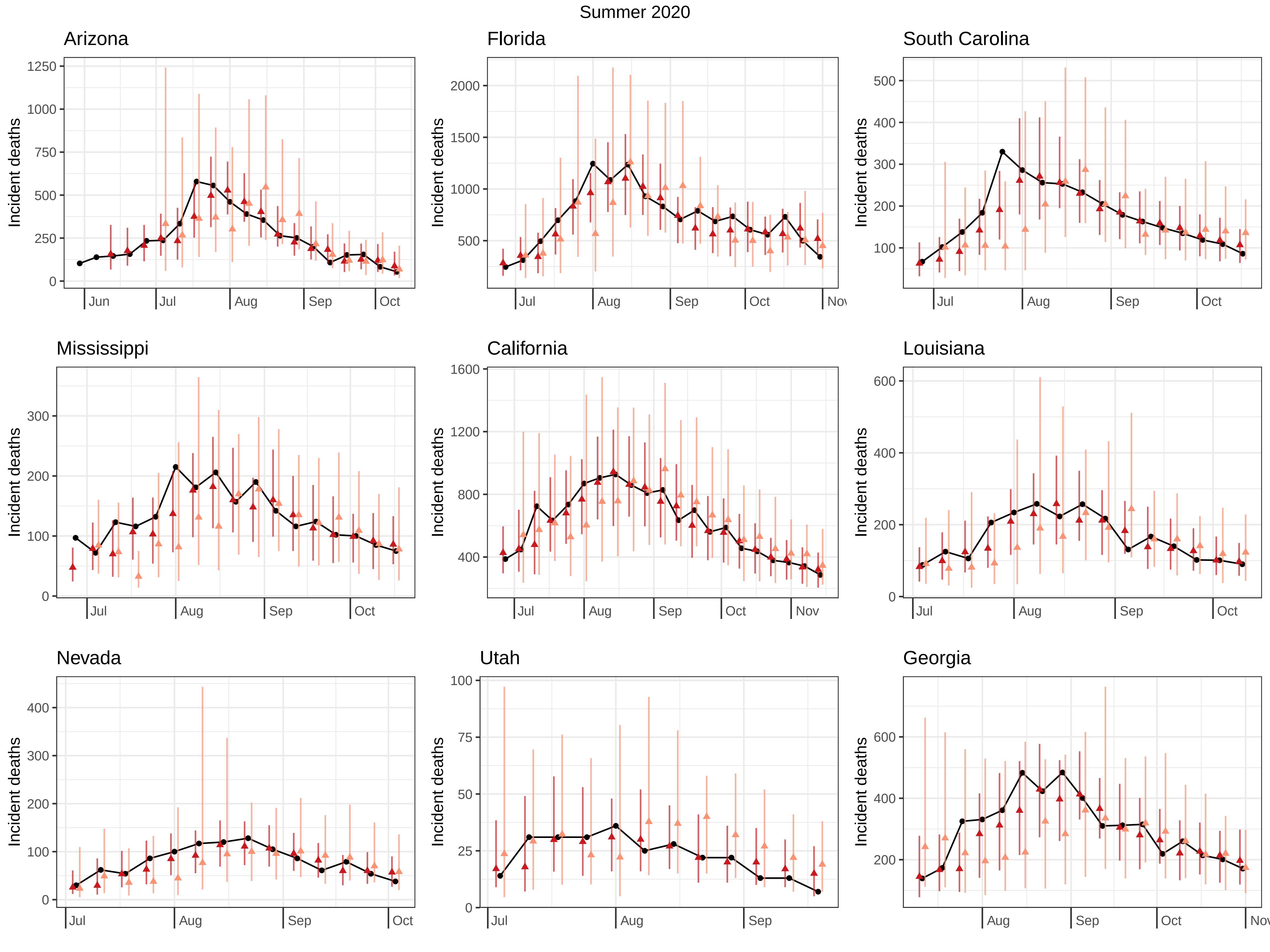

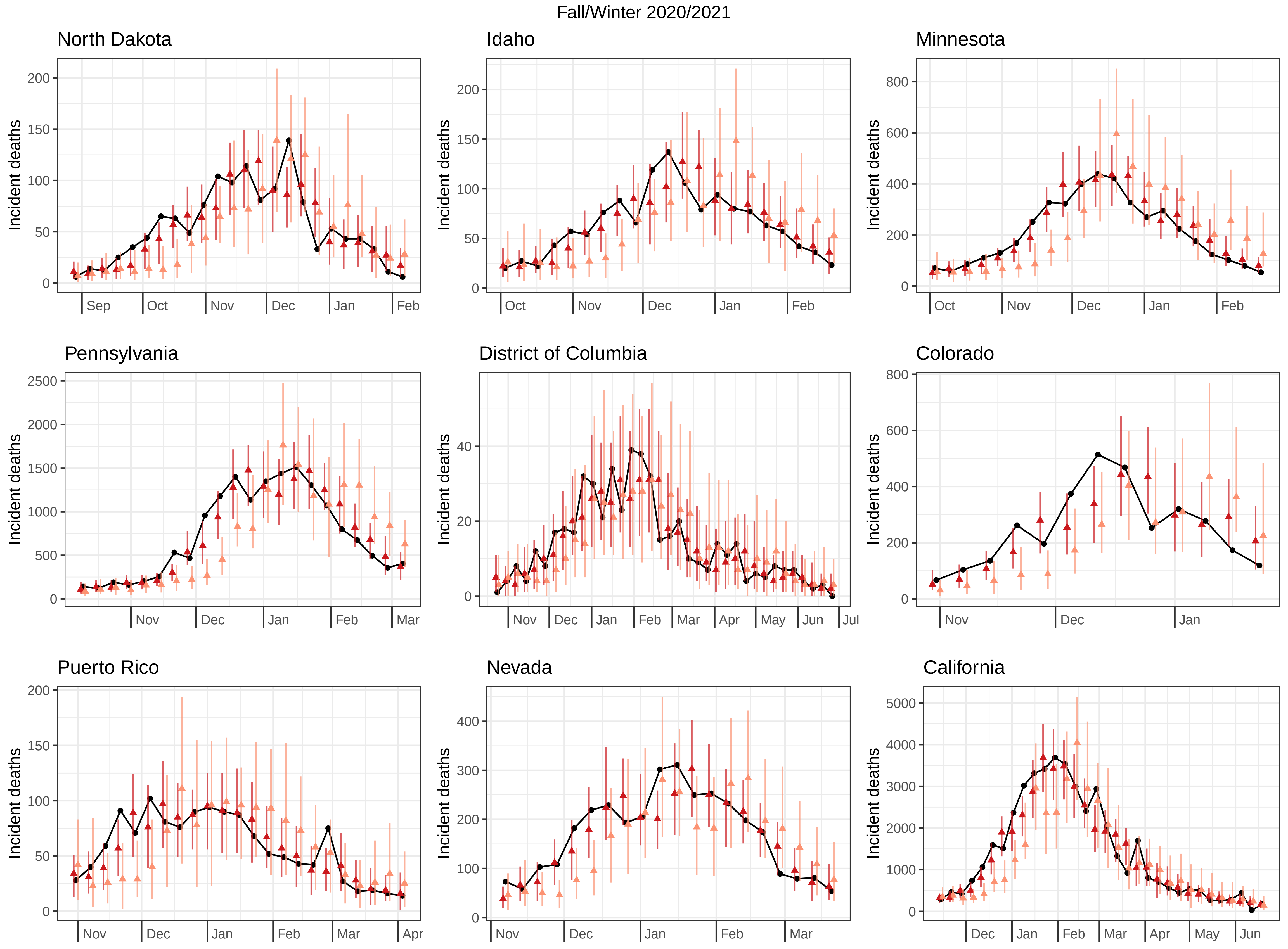

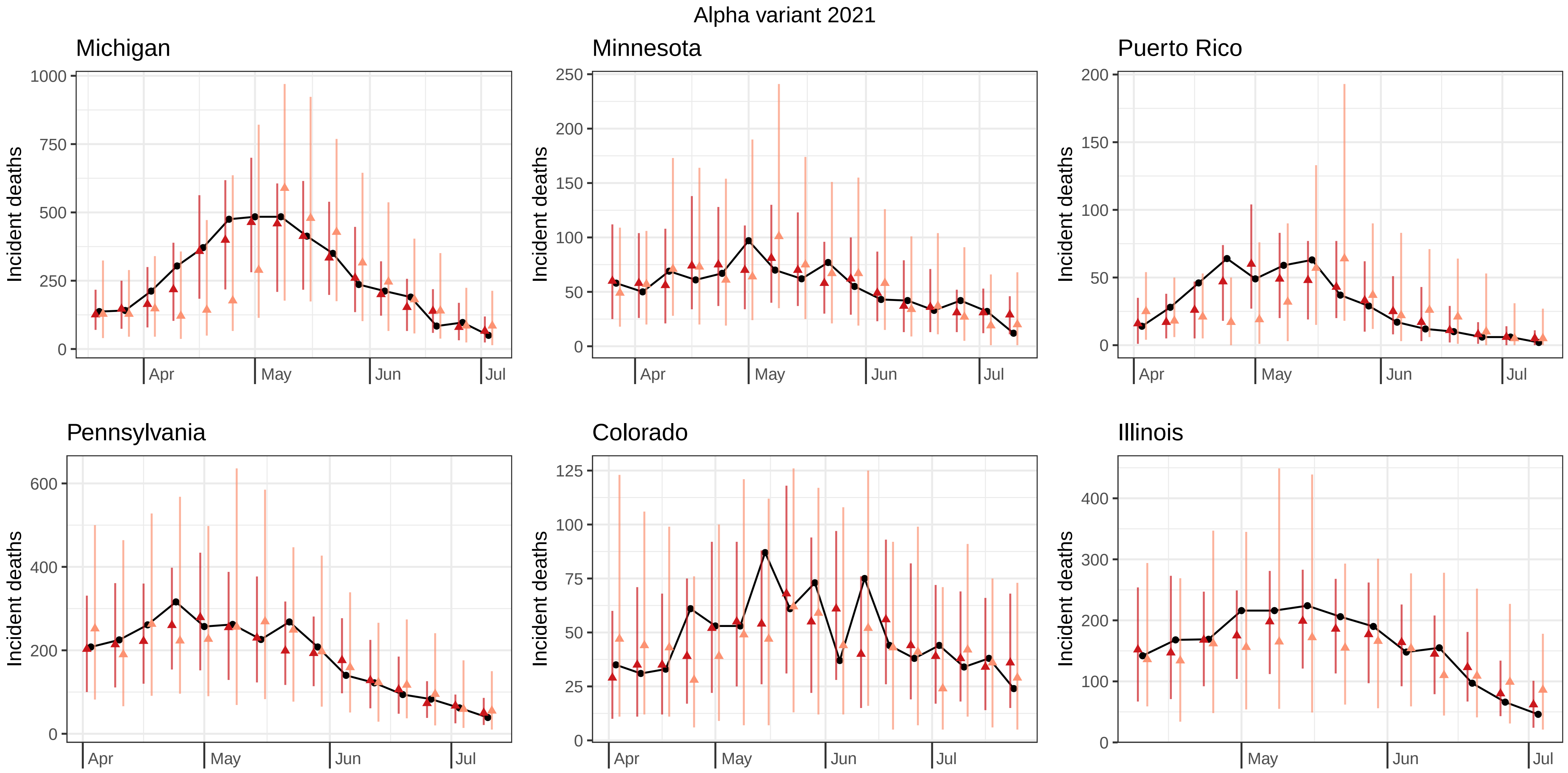

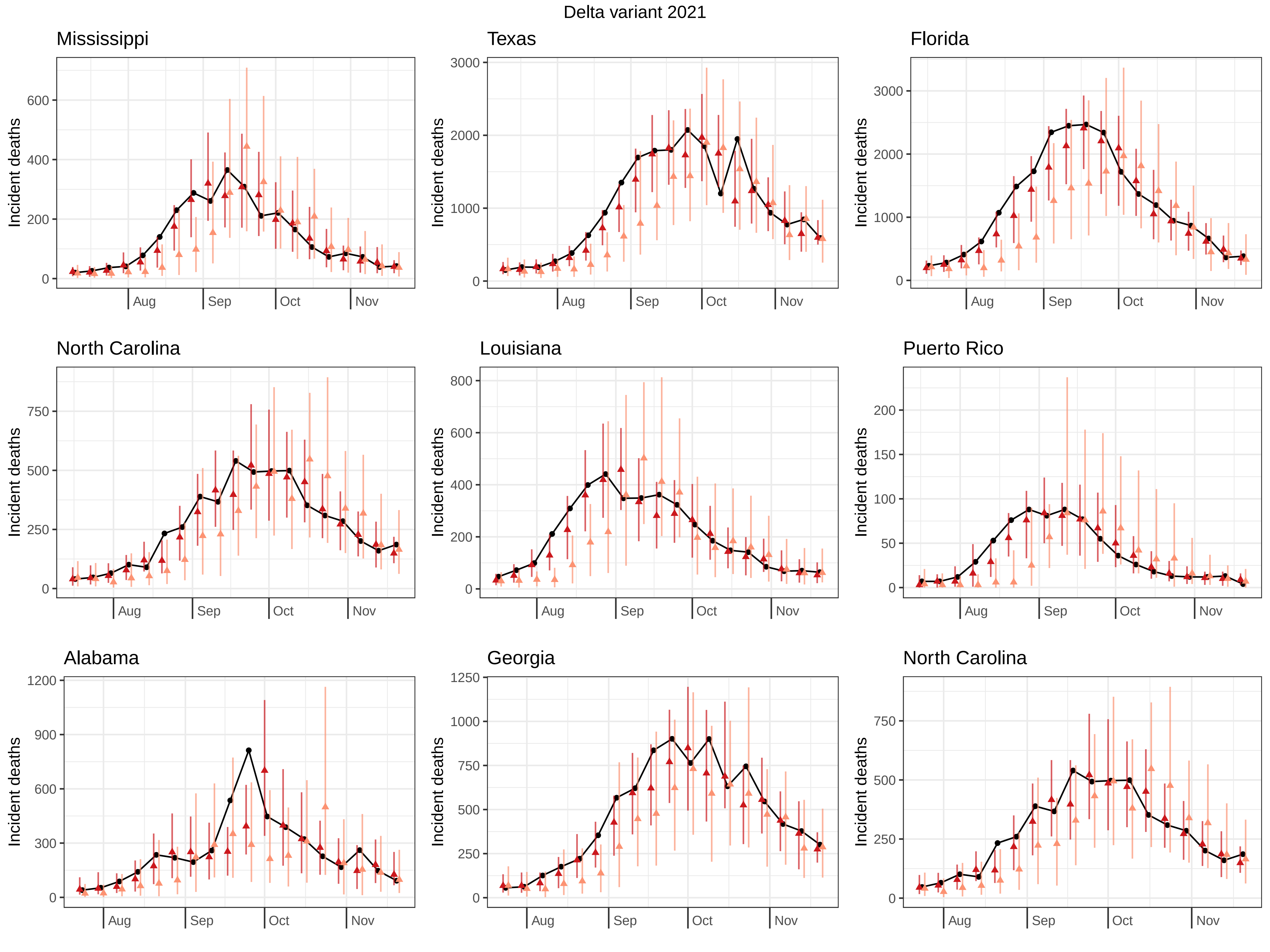
